# Trade-offs between water use and greenhouse gas emissions related to food systems: an optimization study in French adults

**DOI:** 10.1101/2023.03.16.23287343

**Authors:** Emmanuelle Kesse-Guyot, Philippe Pointereau, Joséphine Brunin, Elie Perraud, Hafsa Toujgani, Florine Berthy, Benjamin Allès, Mathilde Touvier, Denis Lairon, François Mariotti, Julia Baudry, Hélène Fouillet

**Affiliations:** Université Sorbonne Paris Nord and Université Paris Cité, Inserm, INRAE, CNAM, Center of Research in Epidemiology and StatisticS (CRESS), Nutritional Epidemiology Research Team (EREN), F-93017 Bobigny, France; Solagro, 75, Voie TOEC, CS 27608, F-31076 Toulouse Cedex 3, France; ADEME, (Agence de l’Environnement et de la Maîtrise de l’Energie), 49004 Angers, France; Université Paris-Saclay, AgroParisTech, INRAE, UMR PNCA, 91120, Palaiseau, France; Aix Marseille Université, Inserm, INRAE, C2VN, 13005 Marseille, France

**Keywords:** GHGe, Water use, compromise modelling, diet optimization, nutrient adequacy, healthy diet

## Abstract

Water use (WU) and greenhouse gas emissions (GHGe) are two main issues facing food systems. Still, they have rarely been studied together even though they are potentially conflicting because their levers for improvement are not necessarily the same. Data on food-related environmental pressures suggest that GHGe and WU can be improved jointly, but their potential conflicts and trade-offs have not been explored. This is what we studied here by a compromise programming approach, using multi-criteria non-linear optimization under a set of nutritional and epidemiological constraints. We used food consumption data of adults aged 18-64 years (n=1,456) from the French representative study INCA 3 (2014-2015) coupled with food environmental impact data from the Agribalyse ® database. A full range of scenarios was identified by prioritizing the two objectives differently, giving weight from 0% to 100%, by 5-% steps, to GHGe improvement over WU improvement.

Overall, we showed that it is possible to achieve significant joint reductions in WU and GHGe relative to their observed values: across the prioritization scenarios, WU reduction ranged from -36% to -14% as its prioritization decreased, while GHGe reduction varied less, from -44 to -52% as its prioritization increased. These joint important reductions in GHGe and WU required the removal of meat consumption (beef, pork, poultry and processed meat), while the consumptions of offal and dairy products remained moderate in order to meet nutrient reference values. However, the consumption of some foods varied according to the priority given to WU over GHGe reductions (namely, vegetables, fruit juice, dairy products, eggs, refined cereal, substitutes, offal and potatoes). Fish, whole grains, and fruit remained more constant due to the epidemiological constraints used. Whatever the scenario, the modeled diets were more plant-based than the observed diet from which they differed significantly (only 23-31% of common food consumptions), and were therefore healthier (63-76% reduction in distance to theoretical minimum risk of chronic disease).

To conclude, while focusing solely on WU reduction induces a joint GHGe reduction that is near-maximal, the reverse is not true, showing that there is good alignment but also some divergence between these objectives. This suggests that food systems WU should be better considered in dietary guidelines for healthy and sustainable diet.

## Introduction

The multiple consequences of climate change include threats to food and water security, deterioration of air quality, climate-related changes in the prevalence of infectious diseases, and overall damage to socio-economic systems (1). Temperature increase by 2100 is estimated at 2.4-3.5°C, and there is about a 50/50 chance that the 1.5°C of the Paris agreement will be exceeded within five years (1). Certain global limits are exceeded for six critical indicators (2–4), including climate change a component of water use (WU), i.e. green WU.

Global food systems based on human activity from field to plate are responsible for approximately a third of total GHGe (5,6). Moreover, the diets of Western countries which are now also widespread in the global South have significant deleterious impacts on human health, with ∼190 million disability-adjusted life years (DALYs) attributed to dietary risk factors worldwide in 2019 (7).

The scientific literature documenting the links between diet and environmental pressures is rich and studies have reported that diets rich in plant-based foods have lower pressures than animal-based foods, notably for GHGe (8–13). While studies on diet-related environmental pressures (14,15) largely focus on climate change and associated GHGe criterion, it is not the only environmental indicator threatened by the food systems, which also significantly impact natural resource depletion and biodiversity loss (4,16,17).

Regarding studies on the water footprint, assessed mainly through blue WU (groundwater or surface water) or green WU (mainly from rainwater and water stored in the soil), the available results are not always consistent and are subject to debate depending on the type of study (water footprint, blue and/or green WU). Indeed, in reviews, healthier diets have been associated with higher (18), similar (in particular for blue water) (19) or lower (8) WU.

Some modeling studies that have alternately optimized different footprints have not reported strong divergences in the composition of diets with lower water and carbon footprints, arguing for synergy rather than conflict between the two environmental indicators (20). However, it has been pointed out that some plant-based foods such as fruits, oils, and nuts are essential components of a healthy diet but are also important contributors to the water footprint and in particular to the blue WU (19,21,22). Thus, while some diet optimization studies have investigated WU (11), an important unresolved question is whether alignments or conflicts (with potential trade-offs) arise between diet-related carbon and water footprints. In addition, no studies have analyzed the similarities or differences between diets that minimize one or the other footprint (18,23,24). Compromise modelling is a key tool to explore such multi-criteria optimization problem, which has not yet been applied to this research question.

In this context, the objective of this study was to use a multicriteria compromise programming approach to analyze the trade-offs between carbon and water footprint reduction by sequentially balancing the minimization of GHGe and WU, under a set of constraints to ensure nutrient security, not worsen long-term health risk, and consider cultural acceptability. This method was applied to the observed diet from a representative French dietary survey combined with the data from the Agribalyse ® database.

## Results

### Description of the sample

The characteristics of the total study population are presented in **Table 1**. The studied population included 1,456 participants (57% women), with a mean age of 42.2 years (SD= 13.5). About 96% of the sample completed three 24-hour recalls.

**Table 1:** Characteristics of study participants, (INCA 3 study, n=1,456)^1^.

|  | **Men** | **Women** |
| --- | --- | --- |
| **N** | 621 | 835 |
| **Age (y)** | 42.05 (13.86) | 42.42 (12.45) |
| **Education** |  |  |
| Primary + College | 42.82 | 40.03 |
| High school | 17.62 | 21.59 |
| Undergraduate level | 19.39 | 20.13 |
| Postgraduate level | 20.03 | 16.54 |
| No information | 0.14 | 1.72 |
| **Physical activity<sup>2</sup> (%)** |  |  |
| Low | 24.4 | 45.56 |
| Moderate | 53.69 | 46.36 |
| High | 21.91 | 8.08 |
| **Living area (%)** |  |  |
| Rural | 27.48 | 23.9 |
| 2,000-19,999 inhab. | 13.91 | 20.04 |
| 20,000-99,999 inhab. | 10.39 | 10.27 |
| ≥100,000 inhab. | 35.88 | 30.97 |
| Paris area | 12.35 | 14.81 |
| **HRS<sup>3</sup> (%)** | 92 (29) | 83 (26) |
| **BMI (kg.m<sup>-2</sup>)** | 25.08 (3.82) | 24.95 (4.94) |
| **Number of 24h recall** | 2.96 (0.19) | 2.95 (0.21) |
| **Energy intake (kcal/d)** | 2682.64 (747.70) | 1963.31 (521.05) |
| **GHGe (kg CO<sub>2</sub> eq /d)** | 6.23 (2.68) | 4.42 (1.50) |
| **WU (m<sup>3</sup> water eq deprivation /d)** | 7.07 (3.76) | 5.82 (3.26) |

Abbreviations: BMI, body mass index; HRS, Health risk score; inhab: inhabitants; WU: water use
<sup>1</sup>Values are n, means (SD) or % as appropriate, all data are weighted on the survey design
<sup>2</sup>Estimated using the RPAQ questionnaire
<sup>3</sup> HRS is the normalized distance to the theoretical minimum-risk exposure levels (TMREL) from the Global Burden of Diseases (i.e. HRS= 0% when the diet is at minimal risk by meeting all the TMREL and HRS=100% when the diet is at maximal risk by deviating from them at most).

### Environmental and other key indicators of modeled diets across trade-off scenarios

Compared to the mean observed diet, the modeled diet issued from the first step, i.e., the one closest to the observed diet meeting the nutritional and epidemiological constraints, had a Diet Similarity Index (DSI, percentage of common food group consumption) of 68%, and 13% higher GHGe (6.05 vs 5.34 kg CO_2_ eq/d), 33% higher WU (8.56 vs 6.46 m^3^ water eq deprivation/d), but 26% lower Health Risk Score (HRS, 64 vs 87 %) (**Supplemental Table 1**).

**Figure 1** and **Supplemental Table 1** depict the environmental and other key indicators (HRS and DSI) for each modeled diet issued from the second step, i.e., from compromise programming by tuning λ from 0% (minimizing WU only) to 100% (minimizing GHGe only) by steps of 5%, always under the nutritional, epidemiological and acceptability constraints. As regards environmental indicators, whatever the trade-off scenario (λ value), GHGe and WU were lower in the modeled than observed diets. From λ = 0% (WU minimization) to 100% (GHGe minimization), GHGe lowered gradually by -14%, from 2.98 to 2.57 kg CO_2_ eq/d (i.e., from -44% to -52% of the observed situation, respectively), while WU increased by 35%, from 4.12 to 5.54 m^3^ water eq deprivation/d (i.e., from - 36% to -14% of the observed situation, respectively). Gradually prioritizing GHGe reduction (from λ=0% to 100%) resulted in diets with a progressive but slight GHGe decrease, while WU showed an initially slight and progressive increase that became more marked from λ≈60%.

**Figure 1:**
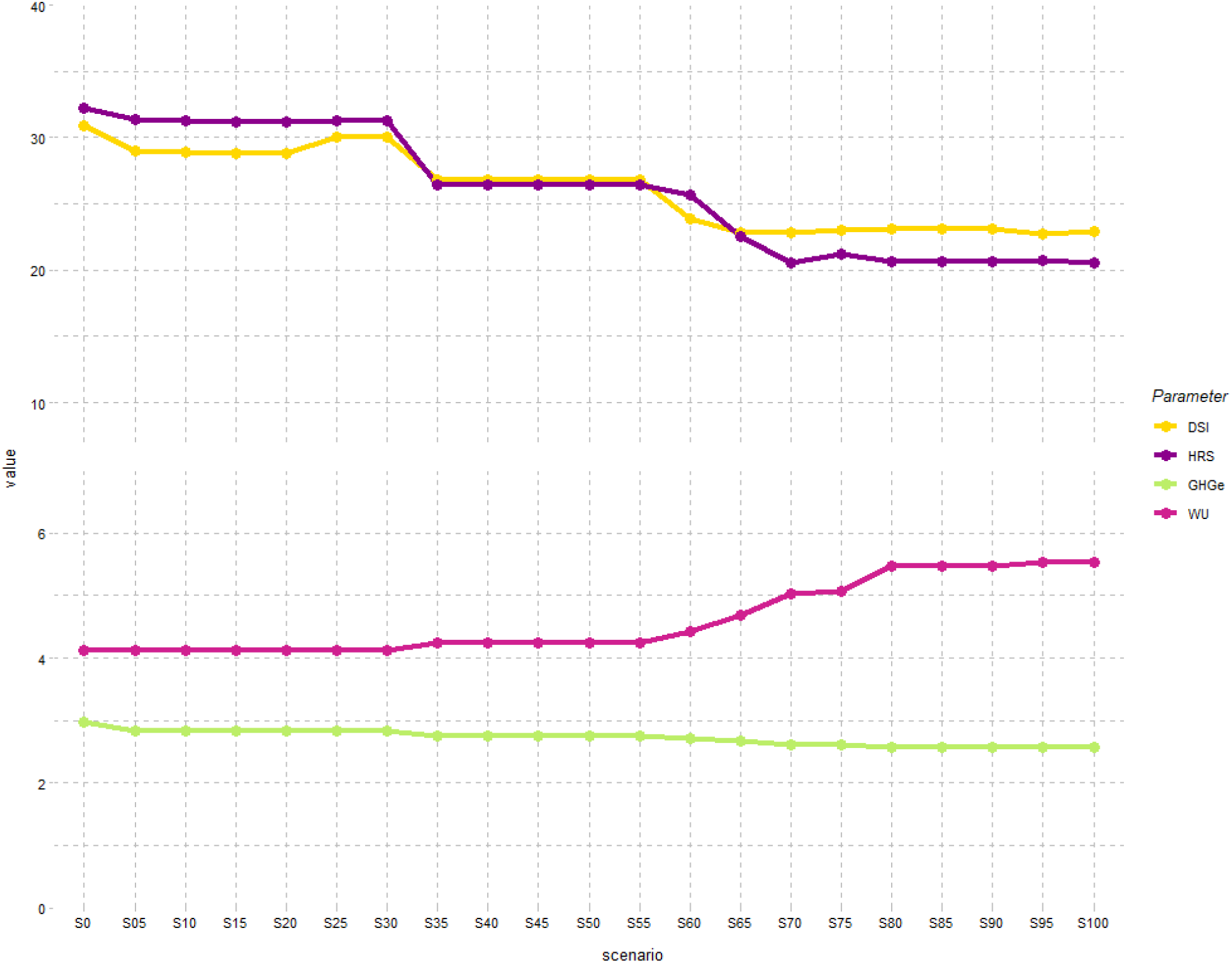
Objective functions and diet descriptors in scenarios differentially prioritizing WU over GHGe minimization^1-3^. Abbreviations: DSI, diet similarity index (%); GHGe, greenhouse gas emissions (kg CO2 eq/d); HRS, health risk score (%); WU, water use (m3 water eq deprivation/d). ^1^ Sx denotes the scenario λ=x% in the compromise programming approach, where λ is the relative weight given to GHGe over WU in the multi-criteria optimization (i.e. S0 and S100 correspond to the minimization of WU only and GHGe only, respectively). ^2^ HRS (%) is the normalized distance to the theoretical minimum-risk exposure levels (TMREL) from the Global Burden of Diseases, expressed in % (i.e., HRS = 0% when the diet is at minimal risk by meeting all the TMREL and HRS=100% when the diet is at maximal risk by deviating from them at most). ^3^ DSI (%) is the proportion of food group consumptions remaining similar to those observed.

Other environmental indicators calculated using the Agribalyse ® database are presented in the **Supplemental Table 2**. Many indicators (notably land use and energy demand) as well as the single aggregated environmental footprint score varied little across the λ range, with similar values at both extreme λ values, but slightly lower values in the middle range (≈-15% for land use, ≈-5% for energy demand and ≈-7% single environmental footprint score for λ=35-60% vs λ=0%).

As regards other key indicators (DSI and HRS, **Figure 1**), all modeled diets were actually greatly distant from the observed diet but also greatly healthier. From λ = 0% to 100%, the modeled diets had no more than 32% to 23% common food group consumptions with the observed diet (DSI) and the estimated health risk (HRS) was decreased to 32% to 21%, respectively (i.e., -63% to -76% decrease compared to the observed situation). DSI and HSR both reached their minimal values for the scenarios with λ ≥ 65% (prioritization of GHGe reduction), but globally they varied only moderately over the λ range.

### Food group consumptions in modeled diets across trade-off scenarios

Compared to the observed diet, some food groups were totally removed from modeled diets, namely beef, processed meat, poultry, and pork, alcoholic beverages, sodas, and other beverages (hot drinks) (**Figure 2** and **Supplemental Table 3**). Across the scenarios gradually prioritizing GHGe reduction (from λ=0% to 100%), there was an increase in eggs, animal fat, legumes, vegetables, and soup, and inversely a decrease in dairy products, offal and snack. Fruit juices were present in the modeled diets up to λ=30% then disappeared. Potatoes, refined cereals, substitutes and dressing showed bell-shaped curves and the other food groups (fish, oil, fruits, nuts and wholegrain products) were present in nearly constant quantities in most of the modeled diets.

**Figure 2:**
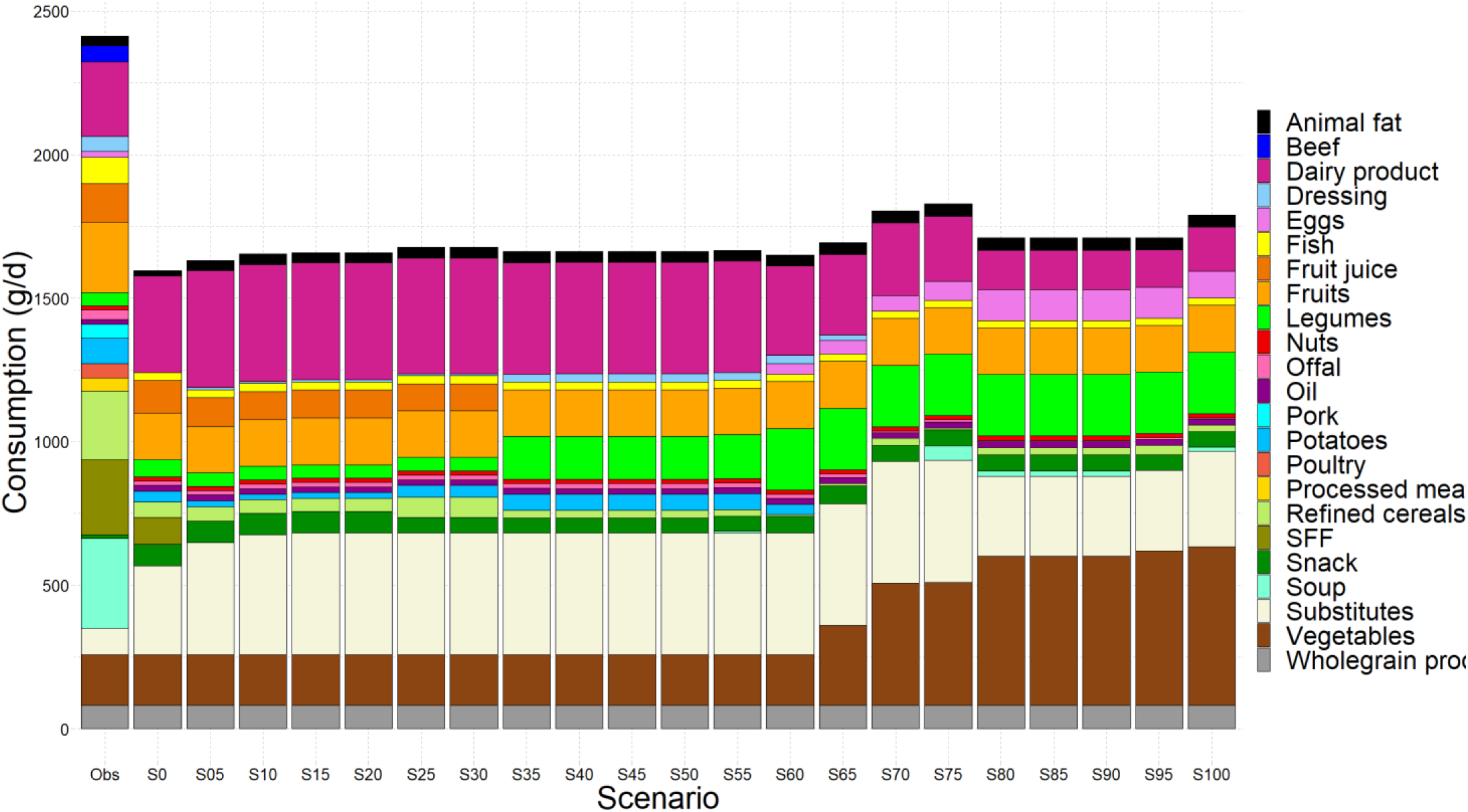
Food group consumptions (g/d) in modeled diets differentially prioritizing WU over GHGe minimization^1^. Abbreviations: GHGe, greenhouse gas emissions; Obs, observed situation; WU: water use. For clarity purpose, the 45 food groups are pooled into 26 broader food categories and beverages are not shown. ^1^ Sx denotes the scenario λ=x% in the compromise programming approach, where λ is the relative weight given to GHGe over WU in the multi-criteria optimization (i.e., S0 and S100 correspond to the minimization of WU only and GHGe only, respectively).

The contributions of food groups to GHGe and WU for each modeled diet are presented in **Supplemental Figure 1 (Supplemental Tables 4 and 5)**. Overall, as meat (beef, pork and poultry) was removed from all modeled diets over the λ range, the main contributors to GHGe were dairy products, offal, eggs, and vegetables and substitutes. The main contributors to WU were vegetables and fruits.

The consumption changes explaining the HRS improvement in the modeled compared to the observed diets were the lower consumptions of sugar-sweetened beverages, red and processed meat and higher consumption of vegetables (**Supplemental Figure 2**). Across the scenarios gradually prioritizing GHGe reduction (from λ=0% to 100%), there was a slight progressive improvement in HRS with the vegetables and legumes increases and red meat reduction.

### Best trade-off and best-balanced diet

The main characteristics and environmental impacts of this diet are presented in **Figure 3** and **Supplemental Table 1**. The best compromise between efficiency and equity in GHGe and WU improvements (i.e., best-balanced solution issued from the last third step) was identified for GHGe of 2.69 kg CO_2_ eq/d and WU of 4.52 m^3^water eq deprivation/d (i.e., -50% and -30% of the observed values, respectively). This best-balanced diet, corresponding to λ≈65%, had a HRS value of 25% (−71% of the observed value) and a DSI of 23%. This diet was composed mainly of dairy products, vegetables, legumes, and substitutes and did not contain any animal flesh, except for a little offal. Some discrepancies were noted between men and women as shown in **Supplemental Figure 3**. Main differences in terms of food group consumptions concerned higher consumption of legumes, offal, nuts, fruits and substitutes in men and higher consumption of vegetables in women.

**Figure 3:**
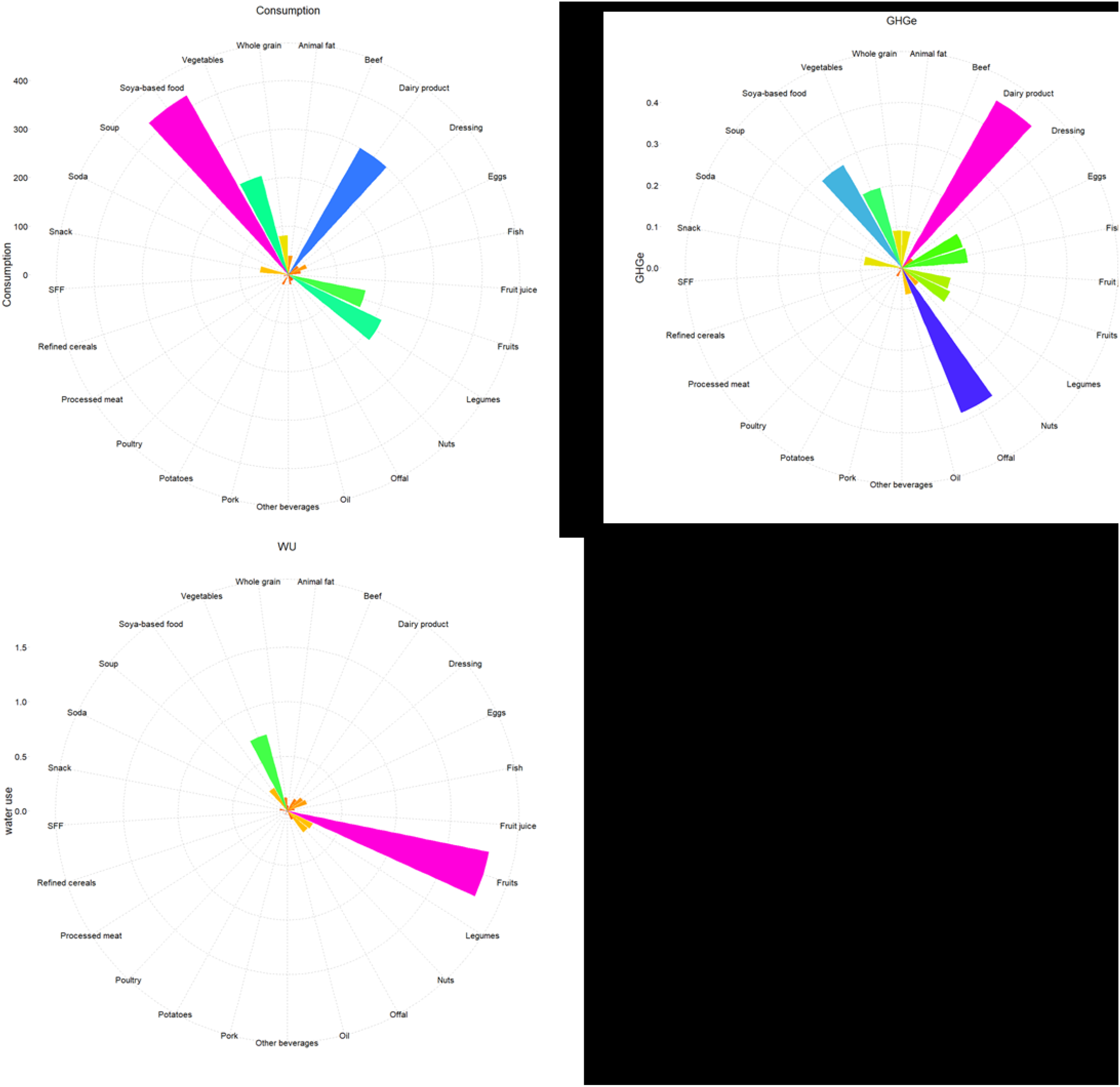
Food group consumptions, GHGe and water use for the best-balanced modeled diet^1^. Abbreviations: GHGe, greenhouse gas emissions; WU: water use. For clarity purpose, the 45 food groups are pooled into 28 broader food categories. ^1^ Food group consumptions in g/d (Panel A,), and contributions of food groups to GHGe in kg CO_2_ eq /d (Panel B,) and WU in m^3^ water eq deprivation /d (Panel C) for the best-balanced modeled diet regarding efficiency and equity, identified by minimizing (d_GHGe+_d_WU_)+max(d_GHGe_, d_WU_), where d_GHGe_ and d_WU_ are the normalized distances to the best GHGe and WU values, respectively.

### Sensitivity analysis

The most limiting nutrients in the modeled diets were quite systematically iodine, sodium, vitamin A, EPA+DHA, and vitamin B2, vitamin C and the linolenic acid / alpha-linoleic acid ratio (data not shown). We tested the influence of being stricter regarding cultural acceptability, by limiting the food group consumptions to their observed 95^th^ rather than 99^th^ centile as in our main analysis. The best-balanced diet identified using these stricter acceptability constraints (**Supplemental Figure 4**) had quite similar WU and GHGe values (4.49 water eq deprivation/d and 2.73 kg CO_2_ eq/d, respectively, Supplemental Table 1), but was characterized by lower consumption of legumes, dairy products and substitutes and higher consumption of refined cereals, potatoes and eggs.

We also tested the influence of being stricter regarding nutritional requirements for bioavailable iron and zinc, by using current reference values rather than lower threshold values ensuring ≤5% deficiency prevalence. The best-balanced diet identified using these stricter nutritional constraints was characterized by higher WU and GHGe values (7.50 water eq deprivation/d and 4.39 kg CO_2_ eq/d, respectively, **Supplemental Table 1**), less dairy product and snack, more fruit juice, eggs, potatoes, sweet and fat foods, and, importantly, the reintroduction of processed meat and poultry that had previously been removed in our main analysis (data not shown).

## Discussion

In the present study we have shown that under a set of nutritional and epidemiological constraints limiting the extent of their potential improvement, it is still possible to reduce WU up to 36% and GHGe up to 52%, and also to reduce them jointly by 30% and 50%, respectively. The modeled diets prioritizing one or the other of these impact reductions share some common main characteristics, such as the absence of some food groups being important contributors to both GHGe and WU (beef, poultry, pork, fatty and sugary products), while they differ in their content in other food groups having more divergent effects on GHGE and WU, such as fruit and vegetables which exhibit low GHGe but elevated WU. The associated human HRS, estimated by the distance to consumptions theoretically ensuring a minimal risk (TMREL), varied little across the modeled diets prioritizing one or the other of these impact reductions. Like the environmental impacts, the health risk was greatly improved compared to the observed situation, at the cost of an important departure from the observed diet (less than 1/3 of unchanged consumption only).

Regarding the nutritional constraints, we considered lowered threshold values than nutritional references for iron and zinc as recently done, allowing for a small increase in anemia to a 5% prevalence (25). This should not be considered a limitation, insofar as we have documented that the current nutritional references for iron and zinc are impediments to identifying healthy and sustainable diets: lowering these thresholds can greatly limit the overall burden of disease as we have previously shown (25), and it also allows greater decreases in environmental pressures. Besides, as expected, and in line with available data in the literature, we found that some nutrients typically provided by animal products are limiting in modeled diets designed to reduce environmental pressure, usually characterized by reduced consumption of animal products (25,26). Furthermore, although the health risk was greatly improved compared to the observed situation, it was still burdened by low overall consumption of wholegrain products. The modeled diets prioritizing GHGe reduction (λ≥0.70) were slightly healthier than others, due to lower consumption of red meat and higher consumption of legumes.

Numerous observational studies have documented that Mediterranean, vegetarian diets with a high contribution of plant-based foods are more favorable to the preservation of natural resources (8,11,15,21,27). Diet optimization studies have also confirmed that reducing the share of animal products in favor of plant-based foods minimizes GHGe (11). For instance, in the NutriNet-Santé study also conducted in France, we have already shown that it is possible to reduce GHGe by 50% from the nutritionally adequate diets by reducing and rearranging the consumptions of animal products while ensuring nutritional adequacy (28). Smaller changes in protein intake in favor of plant protein targeting an increase in nutrient adequacy also decrease GHGe (29).

Although the literature has been growing recently on that topic (19), a lower number of studies have focused on WU in food systems compared to those on GHGs and the footprint indicators used were not consistent (blue, green, or blue and green). Thus, according to a recent review the carbon footprint has been studied twice as much than the water footprint (whatever the indicator used) (30) but the two indicators are rarely studied concomitantly. Furthermore, in diet modeling studies about environmental footprint reductions, environmental indicators were generally included as constraints and not as objectives/targets to be minimized (11), which makes not possible to assess the extent of their maximal improvements and their potential conflicts or alignments. For example, in a study covering 52 countries (31), diets were optimized on different criteria related to sustainability including environmental pressures (GHGe, WU, land use, nitrogen and phosphorus), but introduced as constraints based on planetary boundaries and not as objectives like in the present work. This difference in the methodological approach makes it difficult to compare these results with our own in a meaningful perspective.

In contrast, a diet modeling study that aimed at minimizing WU (in the first step of a hierarchical optimization approach) in food systems was conducted using Hungarian data (32). This study documented a maximum reduction in diet-related water footprint of 19.5% for women and 28.2% for men, these effect sizes being much lower than those we found herein. This may be explained by the use of different indicators (blue and green WU in the quoted study and blue WU herein) and/or by the use of different acceptability constraints, as the Hungarian study more drastically limited the food changes by forcing modeled consumptions to stay between their observed 10^th^ and 90^th^ percentile, which avoided the decline in the animal products being large consumers of green WU (19).

In the available scientific literature, the study with the closest design to our study, offering easier comparison of results, is the study conducted in 2016 by Gephart et al. (20). This study was conducted on US data to identify diets minimizing several environmental footprints (with blue+green water), one by one, under nutritional constraints. The authors concluded that the modeled diets were relatively similar regardless of the optimized indicator (high in plant foods and fish and low in other animal products), and interpreted these results as demonstrating synergies rather than conflicts between environmental indicators.

This is partially consistent with our findings. Indeed, we found that the GHGe and WU values in all the modeled diets, whatever the relative weight given to WU and GHGe in our compromise programming approach, were always markedly lower than their initial values in the observed diets. This argue for a kind of synergy between WU and GHGe in their responses to dietary changes, due to the fact their strongest contributors are often the same food groups (e.g. beef, pork, poultry, offal). However, WU varied much more widely than GHGe over their differential prioritization range. Our results show that the reduction of GHGe is conditional on the reduction of WU, and not vice versa. This is helpful in explaining why some studies have found a moderate decrease or even an increase in WU with low GHGe diets (33). Indeed, our findings illustrate that among low emitting foods (e.g. fresh fruit, vegetables, and some refined and whole cereals), some have quite strong water demands.

In line with our results about the dietary changes required for reducing both WU and GHGe (in particular, those identified in the “*best*” compromise solution) a diet optimization study conducted in India (34) has shown that diets (with constrained energy intake as in our study) should contain less wheat, dairy products, poultry, and nuts, but higher amounts of plants (and especially legumes despite their rather high blue WU) to reduce WU by 18% and 30% by 2025 and 2050 (irrigation water similar to the current value but to feed an increasing population in a 2025 and 2050 scenario), and that these dietary changes should also induce a reduction in GHGe up to -13%. These results seem to concur with ours to indicate that a decrease in water footprint leads to a concomitant GHGe decrease, while the reverse is not systematic according to our results.

Another study used compromise modeling, based on food supply data in Iran, aiming at minimizing WU (blue+green and blue) concomitantly with departure from the original diet under nutritional and import constraints (35). The achieved reductions were 17% and 13% for blue+green and blue only respectively, but unfortunately no data about associated GHGe was provided. However, the dietary changes identified as necessary for WU reduction in this study were consistent with our results, regarding the reduced consumptions of food groups with low “nutritional water productivity” (water demand relative to their nutritional value) such as red meat and poultry, and the increased consumptions (by up to 80% in the Iranian study) of food groups with high “nutritional water productivity”, such as milk, fish, vegetables and legumes, which allowed to comply with certain nutritional references of which animal products are the principal supports. The intensity of the changes in food group consumptions depended on the type of WU to be minimized. Consistent with our model prioritizing WU reduction (blue water in our study), the consumption of food groups with high “nutritional water productivity”, such as fruits and rice, was more strongly affected in the blue water reduction model than in the (blue+green) WU case.

Regarding the nutritional and health dimensions, the modeled diet proposed here as the “best” compromise, which allowed a substantial reduction in both GHGs (−50%) and WU (−30%) compared to the observed situation, had also a reduced human health risk (as estimated using HRS) compared to the observed situation due to the suppression of red meat and increase of plant-based foods (except for wholegrain products, with a consumption remaining far below the TMREL, and limited to the epidemiological constraint (i.e. ≥observed value among consumers). The low consumption of wholegrain products seems to be due to their lack of competitive advantage (or added value) over other food groups: their environmental costs (in terms of GHGe and WU) exceed their nutritional benefits (in terms of nutrients provided).

Also, consumption of substitutes is greatly increased in all optimized models. Therefore, they could be a good lever for water use, climate, and food sustainability. However, if the dietary constraints are more severe (as in the case of iron and zinc), they cannot improve GHGe and water use.

In addition, while these products have lower overall environmental impacts than animal products (36,37), their quality may be compromised because they are often ultra-processed (38). In addition, some adverse health effects of soy foods have been suspected (39).

The best-balanced diet identified here is in line with the optimized diets identified in the literature for GHGe or WU improvements, and characterized by low or no consumption of meat and favoring plant-based foods (8,11). This diet is also globally coherent with the EAT-Lancet diet (21) proposed to preserve both human and planetary health. It should be noted that dairy products and offal (in small quantity) are present in this diet to meet the nutrient reference values, including vitamin A, zinc, iron and calcium.

Our study exhibits some limitations and strengths. First, the major limitation concerns the fact that water footprint considered blue water only. The results are necessarily affected by this limitation because the food groups contributing to blue or green water differ (19). For example, animal products strongly contribute to green water while fruits and cereals strongly contribute to blue water. Second, LCA used herein did not consider the type of farming system (organic or conventional), limiting the consideration of the variety of practices and regionality along the food chain. Data on waste were not available, not allowing to focus on potentially avoidable environmental pressures while some authors have argued that limiting waste throughout the food systems may be an important lever for reducing WU (40). Furthermore, the models use parameters (nutritional contents, nutritional references, TMREL, environmental indicators) which are subject to uncertainties. Finally, optimization applied to the diet has inherent limitations in the methodology, which depends on the options selected in terms of definitions of constraints (e.g., no lower bound for food group consumption in our study), food grouping process and objectives (41). In particular, we considered a detailed foods grouping, which is essential for diet modeling, resulting in some loss of variability in food characteristics, but also avoiding the selection of very specific food items. Concerning strengths, the data were based on LCA according to the standardized guidelines and methodologies environmental data were validated by several expert entities (42). Then, the study was based on diets of a representative sample of French adults, which may allow for generalizability of the findings to other westernized countries. Of note, an innovative compromise programming approach was used to thoroughly describe and understand the potential conflicts between environmental indicators which are multiple and not necessarily in alignment.

## Conclusion

This study is the first to consider the reduction of GHGe and WU in the same optimization model using compromise modelling. While focusing solely on WU reduction induces a joint GHGe reduction that is near-maximal, the reverse is not true, showing that there is good alignment but also some divergence between the two objectives of lowering the carbon and water footprints. Meat is a major contributor to both indicators and the diets limiting both GHGe and WU are much more plant-based than actual diets, but if such more plant-based and healthy diets have low GHGe they may have more or less pronounced WU. Thus, this study suggests that WU in food systems should be better considered to define healthy and sustainable diets, and that otherwise prioritizing a lower-emitting diet per se may be counterproductive in terms of WU.

## Methods

### Population

This study was conducted using data from the INCA 3 study, a nationally representative French survey conducted in 2014-2015 by the French Agency for Food, Environmental and Occupational Health Safety (ANSES). This study initially included 2,121 adult participants who provided food consumption data using a validated method (43). Details of the study design, recruitment and survey plan (definition of individual weight), and methods used have been described in detail elsewhere (43). Overall, participants were selected according to a three-stage random sampling design (geographic units, dwellings, and then individuals) drawn by the National Institute of Statistics and Economic Studies (INSEE). One individual per dwelling was then drawn at random from the eligible individuals at the time of the household contact. The weight of individuals was calculated according to the INSEE method to improve representativeness by region, size of the urban area, occupation and socio-professional category of the household’s reference person, household size, education level, gender, and age (44).

The INCA 3 study protocol was authorized by the National Commission on Informatics and Liberty, after a favorable opinion from the Advisory Committee on Information Processing in Health Research. The study also received a favorable opinion from the Conseil National de l’Information Statistique on 15 June 2011 (n°121/D030) and was awarded the label of “general interest” and statistical quality by the INSEE Label Committee (n°47/Label/D120).

The data collected in the INCA 3 survey encompass food and drink consumption and socio-demographic and lifestyle characteristics. In the present study, we selected adults <65y old (N=1,665) who were not under-reporter (N=1,456) for energy intake (the procedure for identification of under-reporters is described in **Supplemental Method 1**).

### Dietary data

Food and beverage consumption data were collected over 3 non-consecutive days (2 weekdays and 1 weekend day) distributed over approximately three weeks, using the 24-hour recall method by phone conducted by trained interviewers using a standardized validated software for data entry (GloboDiet)(45). Estimation of portion sizes consumed was performed using a picture booklet of food portions and household measurements. Mixed dishes were decomposed in ingredients using the standardized recipes validated by dieticians.

Nutrient intakes were calculated using the 2016 food composition database published by the French Information Centre on Food Quality (46). In the modeling procedure, food consumptions were collapsed into 45 food groups (the list is provided in **Supplemental Table 6**). Nutrient composition and environmental pressure of each food group were calculated as mean over all items of the group weighted by their observed relative consumptions in the food group.

### Health risk and diet similarity scores

Health risk associated with each observed and modeled diet was assessed using the HRS score (33), representing the global normalized distance to the theoretical minimum-risk exposure levels (TMREL) for three unhealthy (red meat, processed meat and sweetened beverages) and six healthy (wholegrain products, fruit, vegetables, legumes, nuts and seeds, and milk) food groups established by the 2019 GBD study (7). The HRS measures the distance to each consumption target (TMREL) weighted by its relative importance using DALYs attributable to each food group in the French population. By construct, HRS varies between 0% and 100%, depending on whether the diet meets all the food group targets (i.e., minimum risk) or deviate from them at most (i.e., maximum risk), respectively. The HRS calculation is presented and detailed in **Supplemental Method 2**.

For each modeled diet, we also computed DSI (diet similarity index) reflecting the proportion of food group consumptions that remained similar to those of the observed diet (47) using the following formula:

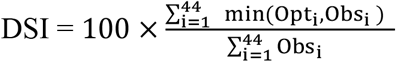

where i denotes the 45 food groups except water used in the optimization model.

### Environmental indicators

Environmental indicators for pressure along with the food chain were estimated using data from the French database Agribalyse ® 3.1 developed by the French Agency for the Environment and Energy Management (ADEME). Agribalyse ® 3.1 contains environmental indicators for 2,517 foods consumed in France. The list was based on the consumption declared in the INCA 3 survey using common coding (48). The methodology has been extensively explained in *ad hoc* published reports (42,49) and is summarized in **Supplemental Method 3**. In the Agribalyse ® database, water footprint has been estimated using the guidelines of The Water Footprint Network (50) and refers to blue water. GHGe and WU for each food group are shown in **Supplemental Table 6**.

### Multicriteria optimization by compromise programming for analyzing GHGe and WU trade-offs

Diet optimization was performed using the procedure SAS/OR ® *optmodel* (version 9.4; SAS Institute, Inc.) using a non-linear optimization algorithm with multi-start option to minimize the risk of obtaining only a local minimum.

Starting from the observed food consumptions, we modeled fully nutrient-adequate diets by including the following constraints in diet optimization:

- Nutritional constraints on daily energy intake and a set of nutrient intakes were based on the recently revised ANSES Reference Values (51) according to the 2021 EFSA opinion (52). For bioavailable iron and zinc, lower bounds were not based on current reference values but on lower threshold values ensuring ≤5% deficiency prevalence, as in our previous study (33), because we have shown that such flexibility enables the identification of healthier diets with a better balance in DALYs due to less cardiometabolic disease, despite a higher prevalence of iron-deficiency anemia (25). Nutritional constraints are presented in **Supplemental Table 7**. For zinc and iron, bioavailability was considered using reference equations (53,54). Details of computation are presented in **Supplemental Method 4**.
- Acceptability constraints were defined by upper bounds set at the weighted 99^th^ percentiles values of each food group (**Supplemental Table 6)**.
- Epidemiological constraints have been also defined to avoid increasing the health risk (as considered by the GBD) beyond its observed level, as follows:
  ∘ at least equal of the observed average (among consumers) consumption for healthy food groups (fruits, vegetables, legumes, nuts, wholegrain products and milk)
  ∘ less than or equal to the observed average intake (among consumers) for the unhealthy food groups (red meat, processed meat and sweetened beverages).

Diet optimization was conducted on the mean dietary data for each sex and for both sexes, by considering an average individual constituted of 50% male and 25% non-menopaused and 25% menopaused female. In the average individual, nutritional references were defined as the weighted values of sex specific nutritional references (**Supplemental Table 1)**, and the 99^th^ and 95^th^ percentiles (see below) of food group consumptions were calculated using the same weighting scheme.

In a first step, we applied the nutritional, epidemiological and acceptability constraints to identify the modifications needed to comply with the nutritional and epidemiologic references only. For this model, the objective function was the minimization of the diet departure (DD) from to the initial (observed) situation using a formula accounting for dietary inertia (55) as:

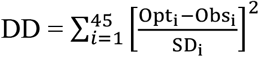

Where Opt_i_ and Obs_i_ denoted the optimized and observed daily consumption of food group (i) and SD_i_ was the standard deviation of the observed daily consumption of food group (i).

Then, the second step consisted in multi-criteria optimization of GHGe and WU by compromise programming. First, we determined the best (minimal) and worst (maximal) values achievable for GHGe and WU, respectively, while satisfying all the model constraints defined above, by mono-criteria optimization as following:

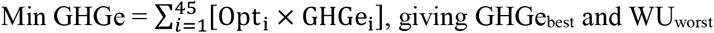

and

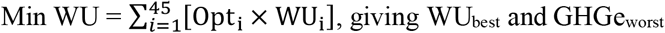

where i is the food group, Opt_i_ denotes the daily consumption of the food group i (g/d) in the optimized model, GHGe_i_ is the greenhouse gas emission for 1 g of the food group i and WU_i_ is the WU for 1 g of the food group i, GHGe_best_ and WU_best_ are the best values and GHGe_worst_ and WU_worst_ the worst values of the corresponding criteria (i.e., the parameters of the pay-off matrix in compromise programming) (26,56).

For purpose of fairness between the GHGe and WU criteria with different units, the multi-criteria optimization was then conducted on the normalized distances to their ideal best values, i.e., on the degree of closeness to ideal points d_GHGe_ and d_WU_ defined by:

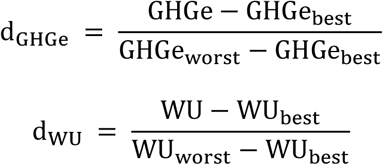

The compromise programming weighted by λand (100% - λ) the d_GHGe_ and d_WU_ terms, respectively, using a multi-objective function (OF) defined as (57,58):

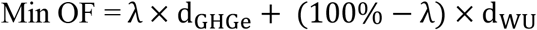

with λ ranging from 0% to 100% by increment of 5%, to explore all the compromise solutions between minimizing only WU (λ =0%) or GHGe (λ =100%).

Finally, in a last third step, we identified the *best-balanced* (BB) solution between the *minsum* (i.e., sum of d_GHGe_ and d_WU_, efficiency metric) and *minmax* (i.e., maximum of d_GHGe_ and d_WU_, equity metric) objectives (26,56,58), as:

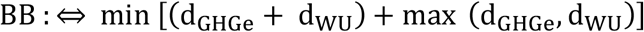

### Sensitivity analysis

First, we analyzed the influence of the acceptability constraints: we used stricter acceptability constraints by lowering the food group consumption limits to their observed 95^th^ percentile, rather than their 99^th^ percentile as in our main analysis.

Second, we analyzed the influence of some nutrient constraints: we used stricter requirements for bioavailable iron and zinc by raising their lower bounds to current reference values (1.92 g/d and 3.62 g/d, respectively), rather than lower threshold values (1.11 g/d and 1.83 g/d, respectively) ensuring ≤5% deficiency prevalence as in our main analysis.

For each modeled diet, we conducted an analysis of the standardized dual values to identify the active and limiting constraints (compared to the inactive ones) that exerted the most influence on the results. The standardized dual values correspond to the potential gain in the objective function in the case of a 100% relaxation of the limiting bound of the constraint (25).

### Statistical analysis

The sociodemographic and lifestyle characteristics of the men and women were presented as mean (SD) or percentage. The observed and modeled diets were described in terms of food group consumptions, environmental pressures, HRS and DSI.

All statistical analyses were performed using SAS® (version 9.4; SAS Institute, Inc., Cary, NC, USA) and figures were drawn using R version 3.6.

## Data Availability

The data collected in the INCA 3 study are available on the website https://www.data.gouv.fr/fr/datasets/donnees-de-consommations-et-habitudes-alimentaires-de-letude-inca-3/
The data from Agribalyse ® are available on the ADEME website: https://agribalyse.ademe.fr/ 

## Abbreviations

DALYs: Disability-Adjusted Life Years
DD: diet departure
DSI: diet similarity index
GBD: Global Burden of Disease
GHGe: Greenhouse gas emissions
HRS: Health Risk Score
INSEE: Institut Nationale des Statistiques et des Etudes Economiques
TMREL: Theoretical Minimum Risk Exposure Level
WU: Water use

## The authors’ contributions

EKG conducted the research, implemented the databases, conducted the analyses and wrote the manuscript; HF provided tools and methodological support; All authors critically helped in the interpretation of results, revised the manuscript and provided relevant intellectual input. They all read and approved the final manuscript; EKG had primary responsibility for the final content, she is the guarantor.

## Data availability

The data collected in the INCA 3 study are available on the website https://www.data.gouv.fr/fr/datasets/donnees-de-consommations-et-habitudes-alimentaires-de-letude-inca-3/

The data from Agribalyse ® are available on the ADEME website: https://agribalyse.ademe.fr/

