## Supplemental materials for "Trade-offs between water use and greenhouse gas emissions related to food systems: an optimization study in French adults"

**Online Supplementary Material**

*Kesse-Guyot et al.*

### Table des matières

**Supplemental Table 1: Environmental and other key indicators in observed and main modeled diets[1-5]**

| | Observed | Closest[1] | Min GHGe (λ=0%)[2] | Min WU (λ=100%)[2] | Best balanced[3] | Best balanced in male[3] | Best balanced in female[3] | Best balanced P95[4] | Best balanced Iron-Zinc[5] |
|---|---|---|---|---|---|---|---|---|---|
| GHGe (kg $CO_2$ eq /d) | 5.34 | 6.05 | 2.98 | 2.57 | 2.69 | 2.87 | 2.41 | 2.73 | 4.39 |
| WU ($m^3$ water eq /d) | 6.46 | 8.56 | 4.12 | 5.54 | 4.52 | 4.54 | 4.16 | 4.49 | 7.50 |
| HRS (%) | 87 | 64 | 32 | 21 | 25 | 27 | 15 | 26 | 30 |
| DSI (%) | - | 68 | 31 | 23 | 23 | 34 | 29 | 34 | 27 |

Abbreviations: DSI, dissimilarity index; GHGe, greenhouse gas emissions; HRS, Health Risk Score; WU, Water Use.

[1] Modeled diet closest to the observed diet while respecting the nutritional, epidemiological and acceptability ($99^{th}$ percentile) constraints (i.e. modeled diet issued from the first step, obtained by minimization of diet departure under constraints).

[2] Modeled diets minimizing GHGe (λ=0%) or WU (λ=100%) while respecting the nutritional, epidemiological and acceptability ($99^{th}$ percentile) constraints.

[3] Best-balanced modeled diet offering the best compromise between GHGe and WU when considering both efficiency and equity, while respecting the nutritional, epidemiological and acceptability ($99^{th}$ percentile) constraints. It was identified in the whole population and also in each sex, by minimizing $(d_{GHGe} + d_{WU}) + \max(d_{GHGe}, d_{WU})$, where $d_{GHGe}$ and $d_{WU}$ are the normalized distances to the best GHGe and WU values, respectively.

[4] Best-balanced modeled diet when using stricter acceptability constraints (upper bounds for food group consumption lowered to their $95^{th}$ percentiles) in sensitivity analysis.

[5] Best-balanced modeled when using stricter nutritional constraints (lower bounds for bioavailable iron and zinc raised to their references values) in sensitivity analysis.

**Supplemental Table 2: Environmental indicators in scenarios differentially prioritizing WU over GHGe minimization[1,2]**

| | S0 | S5 | S10 | S15 | S20 | S25 | S30 | S35 | S40 | S45 | S50 | S55 | S60 | S65 | S70 | S75 | S80 | S85 | S90 | S95 | S100 |
|---|---|---|---|---|---|---|---|---|---|---|---|---|---|---|---|---|---|---|---|---|---|
| Climate change (GHGe, kg $CO_2$ eq) | 2.98 | 2.84 | 2.83 | 2.83 | 2.83 | 2.82 | 2.82 | 2.75 | 2.75 | 2.75 | 2.75 | 2.75 | 2.71 | 2.67 | 2.62 | 2.61 | 2.57 | 2.57 | 2.57 | 2.57 | 2.57 |
| Water use (WU, $m^3$ water eq) | 4.12 | 4.12 | 4.12 | 4.12 | 4.12 | 4.13 | 4.13 | 4.25 | 4.25 | 4.25 | 4.25 | 4.25 | 4.43 | 4.68 | 5.03 | 5.08 | 5.48 | 5.48 | 5.48 | 5.54 | 5.54 |
| Land use (pt) | 37.82 | 37.06 | 37.15 | 37.18 | 37.18 | 36.59 | 36.59 | 32.22 | 32.24 | 32.24 | 32.24 | 32.25 | 34.56 | 35.53 | 36.56 | 37.06 | 38.99 | 38.99 | 38.99 | 39 | 38.6 |
| Energy demand (MJ) | 40.85 | 39.29 | 39.27 | 39.27 | 39.27 | 39.55 | 39.55 | 38.73 | 38.71 | 38.71 | 38.71 | 38.72 | 38.03 | 38.52 | 39.88 | 39.78 | 40.21 | 40.21 | 40.21 | 40.53 | 40.83 |
| Acidification (mol H+ eq) | 0.03 | 0.03 | 0.03 | 0.03 | 0.03 | 0.03 | 0.03 | 0.03 | 0.03 | 0.03 | 0.03 | 0.03 | 0.03 | 0.03 | 0.03 | 0.03 | 0.03 | 0.03 | 0.03 | 0.03 | 0.03 |
| Resource use, minerals and metals (kg Sb eq) | 25.55 | 25.34 | 25 | 24.93 | 24.93 | 24.79 | 24.79 | 23.72 | 23.71 | 23.71 | 23.71 | 23.68 | 23.43 | 23.27 | 22.82 | 22.64 | 23.94 | 23.94 | 23.94 | 24.04 | 23.45 |
| Eutrophication, freshwater (kg P eq) | 0.11 | 0.1 | 0.1 | 0.1 | 0.1 | 0.1 | 0.1 | 0.1 | 0.1 | 0.1 | 0.1 | 0.1 | 0.11 | 0.1 | 0.1 | 0.1 | 0.11 | 0.11 | 0.11 | 0.11 | 0.1 |
| Eutrophication, marine (kg N eq) | 0.53 | 0.51 | 0.51 | 0.51 | 0.51 | 0.5 | 0.5 | 0.49 | 0.49 | 0.49 | 0.49 | 0.49 | 0.51 | 0.52 | 0.52 | 0.52 | 0.54 | 0.54 | 0.54 | 0.54 | 0.54 |
| Eutrophication, terrestrial (mol N eq) | 13.54 | 13.35 | 13.3 | 13.29 | 13.29 | 13.14 | 13.14 | 12.69 | 12.7 | 12.7 | 12.7 | 12.7 | 12.7 | 12.57 | 12.31 | 12.31 | 12.14 | 12.14 | 12.14 | 12.14 | 12.13 |
| Photochemical ozone formation (kg NMVOC eq) | 9.18 | 8.93 | 8.92 | 8.92 | 8.92 | 8.87 | 8.87 | 8.33 | 8.33 | 8.33 | 8.33 | 8.35 | 8.35 | 8.36 | 8.38 | 8.39 | 8.43 | 8.43 | 8.43 | 8.45 | 8.47 |
| Ozone depletion (kg CFC-11eq) | 0.71 | 0.71 | 0.7 | 0.7 | 0.7 | 0.69 | 0.69 | 0.68 | 0.68 | 0.68 | 0.68 | 0.68 | 0.68 | 0.67 | 0.64 | 0.63 | 0.67 | 0.67 | 0.67 | 0.68 | 0.65 |
| Particulate matter (disease incidence) | 0.22 | 0.21 | 0.21 | 0.21 | 0.21 | 0.21 | 0.21 | 0.21 | 0.21 | 0.21 | 0.21 | 0.21 | 0.21 | 0.21 | 0.21 | 0.21 | 0.22 | 0.22 | 0.22 | 0.22 | 0.22 |
| Ionising radiation (kBq U235 eq) | 0.85 | 0.8 | 0.8 | 0.8 | 0.8 | 0.81 | 0.81 | 0.8 | 0.8 | 0.8 | 0.8 | 0.8 | 0.78 | 0.79 | 0.83 | 0.83 | 0.83 | 0.83 | 0.83 | 0.84 | 0.85 |
| Human toxicity, non-cancer [CTUh/person] | 0.07 | 0.06 | 0.06 | 0.06 | 0.06 | 0.06 | 0.06 | 0.06 | 0.06 | 0.06 | 0.06 | 0.06 | 0.06 | 0.06 | 0.06 | 0.06 | 0.06 | 0.06 | 0.06 | 0.06 | 0.06 |
| Human toxicity, cancer [CTUh/person] | 0.24 | 0.23 | 0.23 | 0.23 | 0.23 | 0.23 | 0.23 | 0.22 | 0.22 | 0.22 | 0.22 | 0.23 | 0.24 | 0.24 | 0.25 | 0.25 | 0.26 | 0.26 | 0.26 | 0.26 | 0.26 |
| Ecotoxicity, freshwater [CTUe/person] | 172.13 | 167.95 | 167.59 | 167.59 | 167.59 | 165.23 | 165.23 | 168.56 | 168.6 | 168.6 | 168.6 | 168.59 | 170.95 | 166.18 | 157.86 | 158.11 | 153.41 | 153.41 | 153.41 | 152.91 | 152.4 |
| EF score[4] | 0.37 | 0.36 | 0.36 | 0.36 | 0.36 | 0.36 | 0.36 | 0.35 | 0.35 | 0.35 | 0.35 | 0.35 | 0.36 | 0.36 | 0.36 | 0.36 | 0.37 | 0.37 | 0.37 | 0.37 | 0.37 |

Abbreviations: EF, ecological footprint; GHGe, greenhouse gas emissions; WU, Water Use.

[1]Information about units: kg CO2 eq, carbon dioxide equivalent; m3 world eq, water use in cubic meters of water; land use is estimated as loss of soil organic matter content in kilograms of carbon deficit (kg C deficit) dimensionless and expressed as Points (Pt); MJ, megajoule; mol H+ eq, equivalent of moles hydron; kg Sb eq, equivalent of kilograms of antimony; kg P eq, equivalent of kilograms of phosphorus, kg N eq, equivalent of kilograms of nitrogen; mol N eq, equivalent of moles of nitrogen; kg NMVOC (Non-methane volatile organic compounds) eq, equivalent of kilograms of non-methane volatile organic compounds; kg CFC-11eq, equivalent of kilograms of trichlorofluromethane (Freon-11); Emission of particulate matter in change in mortality due to particulate matter emissions; kg U235 eq, equivalent of kilobecquerels of Uranium 235, CTUh/person, Comparative toxic unit for humans; CTUe/person, ecosystem toxicity unit * volume * time.

[2]Sx denotes the scenario $\lambda$=x% in the compromise programming approach, where $\lambda$ is the relative weight given to GHGe over WU in the multi-criteria optimization (i.e., S0 and S100 correspond to the minimization of WU only and GHGe only, respectively).

**Supplemental Table 3: Food group consumption (g/d) in scenarios differentially prioritizing WU over GHGe minimization[1-3]**

| | Obs | Closest[3] | S0 | S5 | S10 | S15 | S20 | S25 | S30 | S35 | S40 | S45 | S50 | S55 | S60 | S65 | S70 | S75 | S80 | S85 | S90 | S95 | S100 |
|---|---|---|---|---|---|---|---|---|---|---|---|---|---|---|---|---|---|---|---|---|---|---|---|
| Alcoholic beverages | 234 | 0 | 0 | 0 | 0 | 0 | 0 | 0 | 0 | 0 | 0 | 0 | 0 | 0 | 0 | 0 | 0 | 0 | 0 | 0 | 0 | 0 | 0 |
| Animal fat | 10 | 22 | 22 | 35 | 36 | 36 | 36 | 38 | 38 | 38 | 38 | 38 | 38 | 38 | 39 | 41 | 41 | 44 | 44 | 44 | 44 | 42 | 42 |
| Beef | 39 | 0 | 0 | 0 | 0 | 0 | 0 | 0 | 0 | 0 | 0 | 0 | 0 | 0 | 0 | 0 | 0 | 0 | 0 | 0 | 0 | 0 | 0 |
| Dairy products | 162 | 349 | 349 | 407 | 407 | 407 | 407 | 403 | 403 | 389 | 389 | 389 | 389 | 388 | 310 | 281 | 254 | 227 | 137 | 137 | 137 | 130 | 154 |
| Dressing & sauces | 38 | 0 | 0 | 8 | 8 | 8 | 8 | 7 | 7 | 27 | 27 | 27 | 27 | 27 | 29 | 19 | 0 | 0 | 0 | 0 | 0 | 0 | 0 |
| Eggs | 14 | 0 | 0 | 0 | 0 | 0 | 0 | 0 | 0 | 0 | 0 | 0 | 0 | 0 | 38 | 48 | 54 | 65 | 109 | 109 | 109 | 109 | 94 |
| Fish | 32 | 27 | 27 | 27 | 27 | 27 | 27 | 27 | 27 | 27 | 27 | 27 | 27 | 27 | 26 | 26 | 26 | 26 | 24 | 24 | 24 | 24 | 25 |
| Fruit juice | 72 | 110 | 110 | 99 | 99 | 99 | 99 | 94 | 94 | 0 | 0 | 0 | 0 | 0 | 0 | 0 | 0 | 0 | 0 | 0 | 0 | 0 | 0 |
| Fruits | 144 | 162 | 162 | 162 | 162 | 162 | 162 | 162 | 162 | 162 | 162 | 162 | 162 | 162 | 162 | 162 | 162 | 162 | 162 | 162 | 162 | 162 | 162 |
| Legumes | 10 | 47 | 47 | 47 | 47 | 47 | 47 | 47 | 47 | 148 | 150 | 150 | 150 | 153 | 214 | 214 | 214 | 214 | 214 | 214 | 214 | 214 | 214 |
| Nuts | 3 | 15 | 15 | 15 | 15 | 15 | 15 | 15 | 15 | 15 | 15 | 15 | 15 | 15 | 15 | 15 | 15 | 15 | 15 | 15 | 15 | 15 | 15 |
| Offal | 3 | 17 | 17 | 16 | 16 | 16 | 16 | 16 | 16 | 17 | 17 | 17 | 17 | 17 | 15 | 13 | 7 | 8 | 5 | 5 | 5 | 5 | 5 |
| Vegetable oil | 12 | 20 | 20 | 20 | 19 | 19 | 19 | 19 | 19 | 20 | 20 | 20 | 20 | 20 | 20 | 20 | 18 | 20 | 20 | 20 | 20 | 20 | 20 |
| Other beverages | 520 | 0 | 0 | 0 | 0 | 0 | 0 | 0 | 0 | 0 | 0 | 0 | 0 | 0 | 0 | 0 | 0 | 0 | 0 | 0 | 0 | 0 | 0 |
| Pork | 21 | 0 | 0 | 0 | 0 | 0 | 0 | 0 | 0 | 0 | 0 | 0 | 0 | 0 | 0 | 0 | 0 | 0 | 0 | 0 | 0 | 0 | 0 |
| Potatoes | 65 | 0 | 36 | 20 | 19 | 19 | 19 | 43 | 43 | 57 | 56 | 56 | 56 | 55 | 34 | 0 | 0 | 0 | 0 | 0 | 0 | 0 | 0 |
| Poultry | 30 | 36 | 0 | 0 | 0 | 0 | 0 | 0 | 0 | 0 | 0 | 0 | 0 | 0 | 0 | 0 | 0 | 0 | 0 | 0 | 0 | 0 | 0 |
| Processed meat | 40 | 0 | 0 | 0 | 0 | 0 | 0 | 0 | 0 | 0 | 0 | 0 | 0 | 0 | 0 | 0 | 0 | 0 | 0 | 0 | 0 | 0 | 0 |
| Refined cereals | 217 | 0 | 48 | 50 | 47 | 46 | 46 | 70 | 70 | 27 | 26 | 26 | 26 | 23 | 8 | 9 | 24 | 6 | 26 | 26 | 26 | 34 | 23 |
| Sweet and fat foods | 174 | 48 | 75 | 0 | 0 | 0 | 0 | 0 | 0 | 0 | 0 | 0 | 0 | 0 | 0 | 0 | 0 | 0 | 0 | 0 | 0 | 0 | 0 |
| Snack | 3 | 75 | 75 | 75 | 75 | 75 | 75 | 54 | 54 | 52 | 52 | 52 | 52 | 52 | 57 | 61 | 57 | 57 | 57 | 57 | 57 | 55 | 54 |
| Soda | 117 | 75 | 0 | 0 | 0 | 0 | 0 | 0 | 0 | 0 | 0 | 0 | 0 | 0 | 0 | 0 | 0 | 0 | 0 | 0 | 0 | 0 | 0 |
| Soup | 113 | 0 | 0 | 0 | 0 | 0 | 0 | 0 | 0 | 0 | 0 | 0 | 0 | 6 | 0 | 0 | 0 | 50 | 18 | 18 | 18 | 0 | 14 |
| Soya-based food | 4 | 0 | 384 | 391 | 418 | 424 | 424 | 424 | 424 | 424 | 424 | 424 | 424 | 424 | 424 | 424 | 424 | 424 | 278 | 278 | 278 | 279 | 332 |
| Vegetables | 174 | 384 | 176 | 176 | 176 | 176 | 176 | 176 | 176 | 176 | 176 | 176 | 176 | 176 | 176 | 279 | 425 | 429 | 520 | 520 | 520 | 539 | 553 |
| Wholegrain products | 17 | 176 | 82 | 82 | 82 | 82 | 82 | 82 | 82 | 82 | 82 | 82 | 82 | 82 | 82 | 82 | 82 | 82 | 82 | 82 | 82 | 82 | 82 |

Abbreviations: GHGe, greenhouse gas emissions; WU, Water Use.

[1] For clarity purpose, the 45 food groups were collapsed into 26 broader food categories (see Supplemental Table 6 for detailed composition).

[2] Sx denotes the scenario $\lambda$=x% in the compromise programming approach, where $\lambda$ is the relative weight given to GHGe over WU in the multi-criteria optimization (i.e., S0 and S100 correspond to the minimization of WU only and GHGe only, respectively).

[3] Modeled diet closest to the observed diet while respecting the nutritional, epidemiological and acceptability (99th percentile) constraints (i.e. modeled diet issued from the first step, obtained by minimization of diet departure under constraints).

**Supplemental Table 4: Contribution of food groups to GHGe in scenarios differentially prioritizing WU over GHGe minimization[1,2]**

| | S0 | S5 | S10 | S15 | S20 | S25 | S30 | S35 | S40 | S45 | S50 | S55 | S60 | S65 | S70 | S75 | S80 | S85 | S90 | S95 | S100 |
|---|---|---|---|---|---|---|---|---|---|---|---|---|---|---|---|---|---|---|---|---|---|
| Alcoholic beverages | 0.00 | 0.00 | 0.00 | 0.00 | 0.00 | 0.00 | 0.00 | 0.00 | 0.00 | 0.00 | 0.00 | 0.00 | 0.00 | 0.00 | 0.00 | 0.00 | 0.00 | 0.00 | 0.00 | 0.00 | 0.00 |
| Animal fat | 0.04 | 0.06 | 0.06 | 0.06 | 0.06 | 0.06 | 0.06 | 0.06 | 0.07 | 0.07 | 0.07 | 0.06 | 0.09 | 0.09 | 0.11 | 0.09 | 0.09 | 0.09 | 0.09 | 0.07 | 0.07 |
| Beef | 0.00 | 0.00 | 0.00 | 0.00 | 0.00 | 0.00 | 0.00 | 0.00 | 0.00 | 0.00 | 0.00 | 0.00 | 0.00 | 0.00 | 0.00 | 0.00 | 0.00 | 0.00 | 0.00 | 0.00 | 0.00 |
| Dairy products | 0.63 | 0.71 | 0.70 | 0.70 | 0.70 | 0.69 | 0.69 | 0.66 | 0.66 | 0.66 | 0.66 | 0.66 | 0.48 | 0.43 | 0.39 | 0.35 | 0.21 | 0.21 | 0.21 | 0.20 | 0.24 |
| Dressing & sauces | 0.00 | 0.03 | 0.03 | 0.03 | 0.03 | 0.03 | 0.03 | 0.03 | 0.03 | 0.03 | 0.03 | 0.03 | 0.04 | 0.02 | 0.00 | 0.00 | 0.00 | 0.00 | 0.00 | 0.00 | 0.00 |
| Eggs | 0.00 | 0.00 | 0.00 | 0.00 | 0.00 | 0.00 | 0.00 | 0.00 | 0.00 | 0.00 | 0.00 | 0.00 | 0.14 | 0.18 | 0.21 | 0.25 | 0.41 | 0.41 | 0.41 | 0.41 | 0.36 |
| Fish | 0.17 | 0.17 | 0.17 | 0.17 | 0.17 | 0.17 | 0.17 | 0.17 | 0.17 | 0.17 | 0.17 | 0.17 | 0.16 | 0.16 | 0.16 | 0.16 | 0.15 | 0.15 | 0.15 | 0.15 | 0.15 |
| Fruit juice | 0.14 | 0.13 | 0.13 | 0.13 | 0.13 | 0.12 | 0.12 | 0.00 | 0.00 | 0.00 | 0.00 | 0.00 | 0.00 | 0.00 | 0.00 | 0.00 | 0.00 | 0.00 | 0.00 | 0.00 | 0.00 |
| Fruits | 0.12 | 0.12 | 0.12 | 0.12 | 0.12 | 0.12 | 0.12 | 0.12 | 0.12 | 0.12 | 0.12 | 0.12 | 0.12 | 0.12 | 0.12 | 0.12 | 0.12 | 0.12 | 0.12 | 0.12 | 0.12 |
| Legumes | 0.03 | 0.03 | 0.03 | 0.03 | 0.03 | 0.03 | 0.03 | 0.09 | 0.09 | 0.09 | 0.09 | 0.09 | 0.13 | 0.13 | 0.13 | 0.13 | 0.13 | 0.13 | 0.13 | 0.13 | 0.13 |
| Nuts | 0.05 | 0.05 | 0.05 | 0.05 | 0.05 | 0.05 | 0.05 | 0.05 | 0.05 | 0.05 | 0.05 | 0.05 | 0.05 | 0.05 | 0.05 | 0.05 | 0.05 | 0.05 | 0.05 | 0.05 | 0.05 |
| Offal | 0.44 | 0.43 | 0.43 | 0.43 | 0.43 | 0.43 | 0.43 | 0.45 | 0.45 | 0.45 | 0.45 | 0.45 | 0.40 | 0.33 | 0.19 | 0.21 | 0.13 | 0.13 | 0.13 | 0.13 | 0.13 |
| Vegetable oil | 0.07 | 0.07 | 0.06 | 0.06 | 0.06 | 0.06 | 0.06 | 0.07 | 0.07 | 0.07 | 0.07 | 0.07 | 0.07 | 0.07 | 0.06 | 0.07 | 0.07 | 0.07 | 0.07 | 0.07 | 0.07 |
| Other | 0.00 | 0.00 | 0.00 | 0.00 | 0.00 | 0.00 | 0.00 | 0.00 | 0.00 | 0.00 | 0.00 | 0.00 | 0.00 | 0.00 | 0.00 | 0.00 | 0.00 | 0.00 | 0.00 | 0.00 | 0.00 |
| Other beverages | 0.00 | 0.00 | 0.00 | 0.00 | 0.00 | 0.00 | 0.00 | 0.00 | 0.00 | 0.00 | 0.00 | 0.00 | 0.00 | 0.00 | 0.00 | 0.00 | 0.00 | 0.00 | 0.00 | 0.00 | 0.00 |
| Pork | 0.00 | 0.00 | 0.00 | 0.00 | 0.00 | 0.00 | 0.00 | 0.00 | 0.00 | 0.00 | 0.00 | 0.00 | 0.00 | 0.00 | 0.00 | 0.00 | 0.00 | 0.00 | 0.00 | 0.00 | 0.00 |
| Potatoes | 0.04 | 0.02 | 0.02 | 0.02 | 0.02 | 0.04 | 0.04 | 0.06 | 0.06 | 0.06 | 0.06 | 0.05 | 0.03 | 0.00 | 0.00 | 0.00 | 0.00 | 0.00 | 0.00 | 0.00 | 0.00 |
| Poultry | 0.00 | 0.00 | 0.00 | 0.00 | 0.00 | 0.00 | 0.00 | 0.00 | 0.00 | 0.00 | 0.00 | 0.00 | 0.00 | 0.00 | 0.00 | 0.00 | 0.00 | 0.00 | 0.00 | 0.00 | 0.00 |
| Processed meat | 0.00 | 0.00 | 0.00 | 0.00 | 0.00 | 0.00 | 0.00 | 0.00 | 0.00 | 0.00 | 0.00 | 0.00 | 0.00 | 0.00 | 0.00 | 0.00 | 0.00 | 0.00 | 0.00 | 0.00 | 0.00 |
| Refined cereals | 0.04 | 0.04 | 0.04 | 0.04 | 0.04 | 0.06 | 0.06 | 0.02 | 0.02 | 0.02 | 0.02 | 0.02 | 0.01 | 0.01 | 0.02 | 0.00 | 0.02 | 0.02 | 0.02 | 0.03 | 0.02 |
| Sweet and fat foods | 0.24 | 0.00 | 0.00 | 0.00 | 0.00 | 0.00 | 0.00 | 0.00 | 0.00 | 0.00 | 0.00 | 0.00 | 0.00 | 0.00 | 0.00 | 0.00 | 0.00 | 0.00 | 0.00 | 0.00 | 0.00 |
| Snack | 0.12 | 0.12 | 0.12 | 0.12 | 0.12 | 0.09 | 0.09 | 0.08 | 0.08 | 0.08 | 0.08 | 0.08 | 0.09 | 0.10 | 0.09 | 0.09 | 0.09 | 0.09 | 0.09 | 0.09 | 0.08 |
| Soda | 0.00 | 0.00 | 0.00 | 0.00 | 0.00 | 0.00 | 0.00 | 0.00 | 0.00 | 0.00 | 0.00 | 0.00 | 0.00 | 0.00 | 0.00 | 0.00 | 0.00 | 0.00 | 0.00 | 0.00 | 0.00 |
| Soup | 0.00 | 0.00 | 0.00 | 0.00 | 0.00 | 0.00 | 0.00 | 0.00 | 0.00 | 0.00 | 0.00 | 0.01 | 0.00 | 0.00 | 0.00 | 0.01 | 0.00 | 0.00 | 0.00 | 0.00 | 0.00 |
| Soya-based food | 0.26 | 0.26 | 0.28 | 0.29 | 0.29 | 0.29 | 0.29 | 0.29 | 0.29 | 0.29 | 0.29 | 0.29 | 0.29 | 0.29 | 0.29 | 0.29 | 0.19 | 0.19 | 0.19 | 0.19 | 0.22 |
| Vegetables | 0.17 | 0.17 | 0.17 | 0.17 | 0.17 | 0.17 | 0.17 | 0.17 | 0.17 | 0.17 | 0.17 | 0.17 | 0.17 | 0.27 | 0.40 | 0.41 | 0.50 | 0.50 | 0.50 | 0.51 | 0.53 |
| Water | 0.35 | 0.35 | 0.34 | 0.34 | 0.34 | 0.34 | 0.34 | 0.34 | 0.34 | 0.34 | 0.34 | 0.34 | 0.34 | 0.33 | 0.31 | 0.30 | 0.33 | 0.33 | 0.33 | 0.33 | 0.31 |
| Whole grain | 0.09 | 0.09 | 0.09 | 0.09 | 0.09 | 0.09 | 0.09 | 0.09 | 0.09 | 0.09 | 0.09 | 0.09 | 0.09 | 0.09 | 0.09 | 0.09 | 0.09 | 0.09 | 0.09 | 0.09 | 0.09 |

Abbreviations: GHGe, greenhouse gas emissions; WU, Water Use.

[1] For clarity purpose, the 45 food groups are collapsed into 26 broader food categories (see Supplemental Table 6 for detailed composition).

[2] Sx denotes the scenario $\lambda$=x% in the compromise programming approach, where $\lambda$ is the relative weight given to GHGe over WU in the multi-criteria optimization (i.e., S0 and S100 correspond to the minimization of WU only and GHGe only, respectively).

**Supplemental Table 5: Contribution of food group to WU in scenarios differentially prioritizing WU over GHGe minimization [1,2]**

| | S0 | S5 | S10 | S15 | S20 | S25 | S30 | S35 | S40 | S45 | S50 | S55 | S60 | S65 | S70 | S75 | S80 | S85 | S90 | S95 | S100 |
|---|---|---|---|---|---|---|---|---|---|---|---|---|---|---|---|---|---|---|---|---|---|
| Alcoholic beverages | 0.00 | 0.00 | 0.00 | 0.00 | 0.00 | 0.00 | 0.00 | 0.00 | 0.00 | 0.00 | 0.00 | 0.00 | 0.00 | 0.00 | 0.00 | 0.00 | 0.00 | 0.00 | 0.00 | 0.00 | 0.00 |
| Animal fat | 0.02 | 0.04 | 0.04 | 0.04 | 0.04 | 0.04 | 0.04 | 0.04 | 0.04 | 0.04 | 0.04 | 0.04 | 0.05 | 0.05 | 0.06 | 0.05 | 0.05 | 0.05 | 0.05 | 0.05 | 0.05 |
| Beef | 0.00 | 0.00 | 0.00 | 0.00 | 0.00 | 0.00 | 0.00 | 0.00 | 0.00 | 0.00 | 0.00 | 0.00 | 0.00 | 0.00 | 0.00 | 0.00 | 0.00 | 0.00 | 0.00 | 0.00 | 0.00 |
| Dairy products | 0.16 | 0.19 | 0.18 | 0.18 | 0.18 | 0.18 | 0.18 | 0.18 | 0.18 | 0.18 | 0.18 | 0.17 | 0.13 | 0.12 | 0.11 | 0.10 | 0.06 | 0.06 | 0.06 | 0.06 | 0.07 |
| Dressing & sauces | 0.00 | 0.08 | 0.07 | 0.07 | 0.07 | 0.07 | 0.07 | 0.18 | 0.18 | 0.18 | 0.18 | 0.18 | 0.19 | 0.13 | 0.00 | 0.00 | 0.00 | 0.00 | 0.00 | 0.00 | 0.00 |
| Eggs | 0.00 | 0.00 | 0.00 | 0.00 | 0.00 | 0.00 | 0.00 | 0.00 | 0.00 | 0.00 | 0.00 | 0.00 | 0.18 | 0.22 | 0.25 | 0.30 | 0.51 | 0.51 | 0.51 | 0.51 | 0.44 |
| Fish | 0.07 | 0.07 | 0.07 | 0.07 | 0.07 | 0.07 | 0.07 | 0.07 | 0.07 | 0.07 | 0.07 | 0.07 | 0.07 | 0.07 | 0.07 | 0.07 | 0.06 | 0.06 | 0.06 | 0.06 | 0.06 |
| Fruit juice | 0.14 | 0.13 | 0.13 | 0.13 | 0.13 | 0.12 | 0.12 | 0.00 | 0.00 | 0.00 | 0.00 | 0.00 | 0.00 | 0.00 | 0.00 | 0.00 | 0.00 | 0.00 | 0.00 | 0.00 | 0.00 |
| Fruits | 1.89 | 1.89 | 1.89 | 1.89 | 1.89 | 1.89 | 1.89 | 1.89 | 1.89 | 1.89 | 1.89 | 1.89 | 1.89 | 1.89 | 1.89 | 1.89 | 1.89 | 1.89 | 1.89 | 1.89 | 1.89 |
| Legumes | 0.06 | 0.06 | 0.06 | 0.06 | 0.06 | 0.06 | 0.06 | 0.18 | 0.18 | 0.18 | 0.18 | 0.19 | 0.26 | 0.26 | 0.26 | 0.26 | 0.26 | 0.26 | 0.26 | 0.26 | 0.26 |
| Nuts | 0.25 | 0.25 | 0.25 | 0.25 | 0.25 | 0.25 | 0.25 | 0.25 | 0.25 | 0.25 | 0.25 | 0.25 | 0.25 | 0.25 | 0.25 | 0.25 | 0.25 | 0.25 | 0.25 | 0.25 | 0.25 |
| Offal | 0.10 | 0.10 | 0.10 | 0.10 | 0.10 | 0.10 | 0.10 | 0.11 | 0.10 | 0.10 | 0.10 | 0.11 | 0.09 | 0.08 | 0.04 | 0.05 | 0.03 | 0.03 | 0.03 | 0.03 | 0.03 |
| Vegetable oil | 0.01 | 0.01 | 0.01 | 0.01 | 0.01 | 0.01 | 0.01 | 0.01 | 0.01 | 0.01 | 0.01 | 0.01 | 0.01 | 0.01 | 0.01 | 0.01 | 0.01 | 0.01 | 0.01 | 0.01 | 0.01 |
| Other | 0.00 | 0.00 | 0.00 | 0.00 | 0.00 | 0.00 | 0.00 | 0.00 | 0.00 | 0.00 | 0.00 | 0.00 | 0.00 | 0.00 | 0.00 | 0.00 | 0.00 | 0.00 | 0.00 | 0.00 | 0.00 |
| Other beverages | 0.00 | 0.00 | 0.00 | 0.00 | 0.00 | 0.00 | 0.00 | 0.00 | 0.00 | 0.00 | 0.00 | 0.00 | 0.00 | 0.00 | 0.00 | 0.00 | 0.00 | 0.00 | 0.00 | 0.00 | 0.00 |
| Pork | 0.00 | 0.00 | 0.00 | 0.00 | 0.00 | 0.00 | 0.00 | 0.00 | 0.00 | 0.00 | 0.00 | 0.00 | 0.00 | 0.00 | 0.00 | 0.00 | 0.00 | 0.00 | 0.00 | 0.00 | 0.00 |
| Potatoes | 0.05 | 0.03 | 0.03 | 0.03 | 0.03 | 0.06 | 0.06 | 0.08 | 0.08 | 0.08 | 0.08 | 0.07 | 0.05 | 0.00 | 0.00 | 0.00 | 0.00 | 0.00 | 0.00 | 0.00 | 0.00 |
| Poultry | 0.00 | 0.00 | 0.00 | 0.00 | 0.00 | 0.00 | 0.00 | 0.00 | 0.00 | 0.00 | 0.00 | 0.00 | 0.00 | 0.00 | 0.00 | 0.00 | 0.00 | 0.00 | 0.00 | 0.00 | 0.00 |
| Processed meat | 0.00 | 0.00 | 0.00 | 0.00 | 0.00 | 0.00 | 0.00 | 0.00 | 0.00 | 0.00 | 0.00 | 0.00 | 0.00 | 0.00 | 0.00 | 0.00 | 0.00 | 0.00 | 0.00 | 0.00 | 0.00 |
| Refined cereals | 0.02 | 0.03 | 0.02 | 0.02 | 0.02 | 0.04 | 0.04 | 0.01 | 0.01 | 0.01 | 0.01 | 0.01 | 0.00 | 0.00 | 0.01 | 0.00 | 0.01 | 0.01 | 0.01 | 0.02 | 0.01 |
| Sweet and fat foods | 0.08 | 0.00 | 0.00 | 0.00 | 0.00 | 0.00 | 0.00 | 0.00 | 0.00 | 0.00 | 0.00 | 0.00 | 0.00 | 0.00 | 0.00 | 0.00 | 0.00 | 0.00 | 0.00 | 0.00 | 0.00 |
| Snack | 0.09 | 0.09 | 0.09 | 0.09 | 0.09 | 0.06 | 0.06 | 0.06 | 0.06 | 0.06 | 0.06 | 0.06 | 0.07 | 0.07 | 0.07 | 0.07 | 0.07 | 0.07 | 0.07 | 0.06 | 0.06 |
| Soda | 0.00 | 0.00 | 0.00 | 0.00 | 0.00 | 0.00 | 0.00 | 0.00 | 0.00 | 0.00 | 0.00 | 0.00 | 0.00 | 0.00 | 0.00 | 0.00 | 0.00 | 0.00 | 0.00 | 0.00 | 0.00 |
| Soup | 0.00 | 0.00 | 0.00 | 0.00 | 0.00 | 0.00 | 0.00 | 0.00 | 0.00 | 0.00 | 0.00 | 0.01 | 0.00 | 0.00 | 0.00 | 0.00 | 0.00 | 0.00 | 0.00 | 0.00 | 0.00 |
| Soya-based food | 0.22 | 0.22 | 0.24 | 0.24 | 0.24 | 0.24 | 0.24 | 0.24 | 0.24 | 0.24 | 0.24 | 0.24 | 0.24 | 0.24 | 0.24 | 0.24 | 0.16 | 0.16 | 0.16 | 0.16 | 0.19 |
| Vegetables | 0.61 | 0.61 | 0.61 | 0.61 | 0.61 | 0.61 | 0.61 | 0.61 | 0.61 | 0.61 | 0.61 | 0.61 | 0.61 | 0.96 | 1.46 | 1.47 | 1.79 | 1.79 | 1.79 | 1.85 | 1.90 |
| Water | 0.23 | 0.23 | 0.22 | 0.22 | 0.22 | 0.22 | 0.22 | 0.22 | 0.22 | 0.22 | 0.22 | 0.22 | 0.22 | 0.22 | 0.20 | 0.19 | 0.21 | 0.21 | 0.21 | 0.21 | 0.20 |
| Whole grain | 0.12 | 0.12 | 0.12 | 0.12 | 0.12 | 0.12 | 0.12 | 0.12 | 0.12 | 0.12 | 0.12 | 0.12 | 0.12 | 0.12 | 0.12 | 0.12 | 0.12 | 0.12 | 0.12 | 0.12 | 0.12 |

Abbreviations: GHGe, greenhouse gas emissions; WU, Water Use.

[1] For clarity purpose, the 45 food groups were collapsed into 26 broader food categories (see Supplemental Table 6 for detailed composition)

[2] Sx denotes the scenario λ=x% in the compromise programming approach, where λ is the relative weight given to GHGe over WU in the multi-criteria optimization (i.e., S0 and S100 correspond to the minimization of WU only and GHGe only, respectively).

**Supplemental Table 6: Main characteristics of the 45 food groups and 28 food categories included in the diet optimization models[1]**

| Food category | Food group | Number of food items | Number of consumers | Mean consumption[1] | 95th percentile of the distribution[1] | 99th percentile of the distribution[1] | Greenhouse gas emissions (kg $CO_2$ eq/100g) | Blue Water footprint ($m^3$ world eq/100g) |
|---|---|---|---|---|---|---|---|---|
| Alcoholic beverages | Alcoholic beverages | 34 | 951 | 234 | 817 | 1237 | 0.112 | 0.061 |
| Animal fat | Animal fats | 4 | 36 | 20 | 64 | 71 | 0.163 | 0.111 |
| | Low fat butters and buttermilk | 11 | 1148 | 12 | 34 | 46 | 0.782 | 0.25 |
| Beef | Beef and veal | 38 | 916 | 56 | 174 | 263 | 3.628 | 0.8 |
| Dairy product | Milk | 14 | 937 | 128 | 428 | 755 | 0.155 | 0.043 |
| | Dairy products | 18 | 639 | 86 | 214 | 286 | 0.215 | 0.051 |
| | Cheese | 93 | 1373 | 46 | 106 | 153 | 0.556 | 0.216 |
| Dressing | Salt | 5 | 1331 | 2 | 4 | 6 | 0.061 | 0.041 |
| | Condiments | 10 | 379 | 12 | 43 | 55 | 0.127 | 0.673 |
| | Herbs Spices | 37 | 1249 | 2 | 7 | 13 | 0.36 | 0.916 |
| | Fresh sauces and creams | 51 | 1346 | 35 | 103 | 150 | 0.428 | 0.338 |
| Eggs | Eggs | 13 | 1002 | 22 | 70 | 109 | 0.38 | 0.464 |
| Fish | Fatty fish | 29 | 479 | 26 | 77 | 126 | 0.614 | 0.255 |
| | Lean fish | 54 | 633 | 46 | 127 | 180 | 1.161 | 0.453 |
| | Mollusks | 15 | 364 | 18 | 95 | 118 | 1.41 | 0.237 |
| Fruit Juice | Fruit juices | 26 | 797 | 137 | 357 | 598 | 0.127 | 0.131 |
| Fruits | Fresh fruits | 49 | 1177 | 162 | 416 | 585 | 0.075 | 1.162 |
| | Dried fruits | 9 | 156 | 16 | 74 | 146 | 0.201 | 4.587 |
| | Processed fruits | 11 | 348 | 65 | 165 | 236 | 0.094 | 0.326 |
| Legumes | Pulses | 15 | 263 | 47 | 146 | 214 | 0.061 | 0.122 |
| Nuts | Nuts | 23 | 335 | 15 | 45 | 64 | 0.363 | 1.676 |
| Offal | Offal | 17 | 100 | 33 | 73 | 107 | 2.62 | 0.613 |
| Oil | ALA-rich vegetable fats | 3 | 130 | 4 | 13 | 20 | 0.324 | 0.048 |

| | | | | | | | | |
|---|---|---|---|---|---|---|---|---|
| | ALA-poor vegetable oils and fats | 25 | 1368 | 12 | 30 | 59 | 0.374 | 1.073 |
| Other | Other foods | 11 | 704 | 6 | 27 | 43 | 0.146 | 0.228 |
| Other beverages | Hot drinks | 22 | 1399 | 545 | 1210 | 1840 | 0.062 | 0.05 |
| Pork | Pork | 30 | 622 | 49 | 131 | 265 | 1.562 | 0.955 |
| Potatoes | Potatoes | 17 | 1039 | 89 | 250 | 577 | 0.1 | 0.136 |
| Poultry | Poultry | 24 | 870 | 50 | 119 | 171 | 0.897 | 0.879 |
| Processed meat | Processed meat | 62 | 1275 | 46 | 131 | 194 | 1.03 | 0.587 |
| Refined cereals | Refined bread and bakery products | 33 | 1418 | 141 | 334 | 470 | 0.079 | 0.052 |
| | Other refined cereals | 12 | 1196 | 98 | 243 | 370 | 0.106 | 1.274 |
| Snack | Salty/fatty products | 15 | 344 | 13 | 43 | 75 | 0.157 | 0.115 |
| Soda | Sweetened drinks | 40 | 674 | 250 | 794 | 1684 | 0.047 | 0.143 |
| Soup | Soups | 30 | 439 | 285 | 634 | 1082 | 0.112 | 0.117 |
| | Broths | 7 | 243 | 29 | 111 | 281 | 0.014 | 0.01 |
| Substitutes | Substitutes | 9 | 101 | 91 | 247 | 424 | 0.067 | 0.057 |
| Sweet and/or fat products | Sweet/fat products | 57 | 675 | 40 | 103 | 160 | 0.309 | 0.54 |
| | Sweetened dairy products | 37 | 548 | 87 | 205 | 304 | 0.185 | 0.135 |
| | Sweetened milk desserts | 21 | 805 | 43 | 114 | 196 | 0.322 | 0.101 |
| | Sweet and/or fatty products | 185 | 1433 | 91 | 228 | 340 | 0.406 | 0.388 |
| Vegetables | Vegetables | 148 | 1441 | 176 | 409 | 553 | 0.095 | 0.344 |
| Water | Water | 43 | 1431 | 965 | 2175 | 3121 | 0.027 | 0.017 |
| Wholegrain products | Wholemeal bread and bakery products | 15 | 522 | 43 | 116 | 206 | 0.112 | 0.15 |
| | Other whole grains | 11 | 153 | 39 | 95 | 149 | 0.128 | 0.704 |

[1]based on the weighted distribution among consumers in the INCA 3 study

[2] constituted of soy beverage (3/9), almond drink (1/9), soy dessert (3/9), tofu (1/9) and soy patty or steak (1/9)

**Supplemental Table 7: Nutritional constraints used in the optimization models**

| | Male | | Female | | Average individual[1] | |
|---|---|---|---|---|---|---|
| | **Lower reference** | **Upper reference** | **Lower reference** | **Upper reference** | **Lower reference** | **Upper reference** |
| Energy intake | ER - 5% | ER - 5% | ER - 5% | ER - 5% | ER - 5% | ER - 5% |
| Protein | 0.83 × bw | 2.3 × bw | 0.83 × bw | 2.3 × bw | 0.83 × bw | 2.3 × bw |
| Vitamin A | 750 µg | 3000 µg | 650 µg | 3000 µg | 700 µg | 3000 µg |
| Vitamin B1 | 0.42 µg /kcal | - | 0.42µg /kcal | - | 0.42µg /kcal | - |
| Vitamin B2 | 1.60 mg | - | 1.60 mg | - | 1.60 mg | - |
| Vitamin B3 | 6.7 µg /kcal | 900 µg | 6.7 µg /kcal | 900 µg | 6.7 µg /kcal | 900 µg |
| Vitamin B5 | P5 mg | - | P5 mg | - | Weighted P5 | - |
| Vitamin B6 | 1.7 mg | 25 mg | 1.6 mg | 25 mg | 1.65 mg | 25 mg |
| Vitamin B9 | 330 µg | - | 330 µg | - | 330 µg | - |
| Vitamin B12 | 4 µg | - | 4 µg | - | 4 µg | - |
| Vitamin C | 110 mg | - | 110 mg | - | 110 mg | - |
| Vitamin E | P5 g | - | P5 g | - | Weighted P5 | - |
| Vitamin K | P5 µg | - | P5 µg | - | Weighted P5 | - |
| Calcium | 950 g | 2500 g | 950 g | 2500 g | 950 g | 2500 g |
| Copper | P5 g | 5 g | P5 g | 5 g | Weighted P5 | 5 g |
| Bioavailable Iron | 1.10 mg | - | 1.10/ 1.16 mg (M-)/(M+) | - | 1.115 mg | - |
| Iodine | 150 µg | 600 µg | 150 µg | 600 µg | 150 µg | 600 µg |
| Magnesium | P5 g | - | P5 g | - | Weighted P5 | - |
| Manganese | P5 g | - | P5 g | - | Weighted P5 | - |
| Phosphorus | 550 mg | - | 550 mg | - | 550 mg | - |
| Potassium | 3500 mg | - | 3500 mg | - | 3500 mg | - |
| Selenium | 70 µg | 300 µg | 70 µg | 300 µg | 70 µg | 300 µg |
| Sodium | 1500 mg | 2300 mg | 1500 mg | 2300 mg | 1500 mg | 2300 mg |
| Bioavailable zinc | 2.06 mg | | 1.61 mg | | 1.835 mg | |
| SFA | - | 12% EI | - | 12% EI | - | 12% EI |
| Linoleic acid | 4% EI | - | 4% EI | - | 4% EI | - |
| ALA | 1% EI | - | 1% EI | - | 1% EI | - |
| LA / ALA | - | 5 | - | 5 | - | 5 |
| EPA+DHA | 0.5 g | - | 0.5 g | - | 0.5 g | - |
| Sugar without lactose | - | 100 g | - | 100 g | - | 100 g |
| Fiber | 30 g | - | 30 g | - | 30 g | - |

Abbreviations: ALA, alpha-linoleic acid; bw, body weight (kg); DHA, docosahexaenoic acid; EI, energy intake; EPA, eicosapentaenoic acid; ER, energy requirement; LA, linolenic acid; $M^-$, non-menopausal; $M^+$, menopausal; SFA, saturated fatty acids

[1] In case of different references between men and women. The average individual was the weighted mean as follows: 50% men, 25% women $M^-$, 25% $M^+$.

**Supplemental Figure 1: Greenhouse gas emissions and water use from food groups across water use-greenhouse gas emissions trade-off models[1,2]**

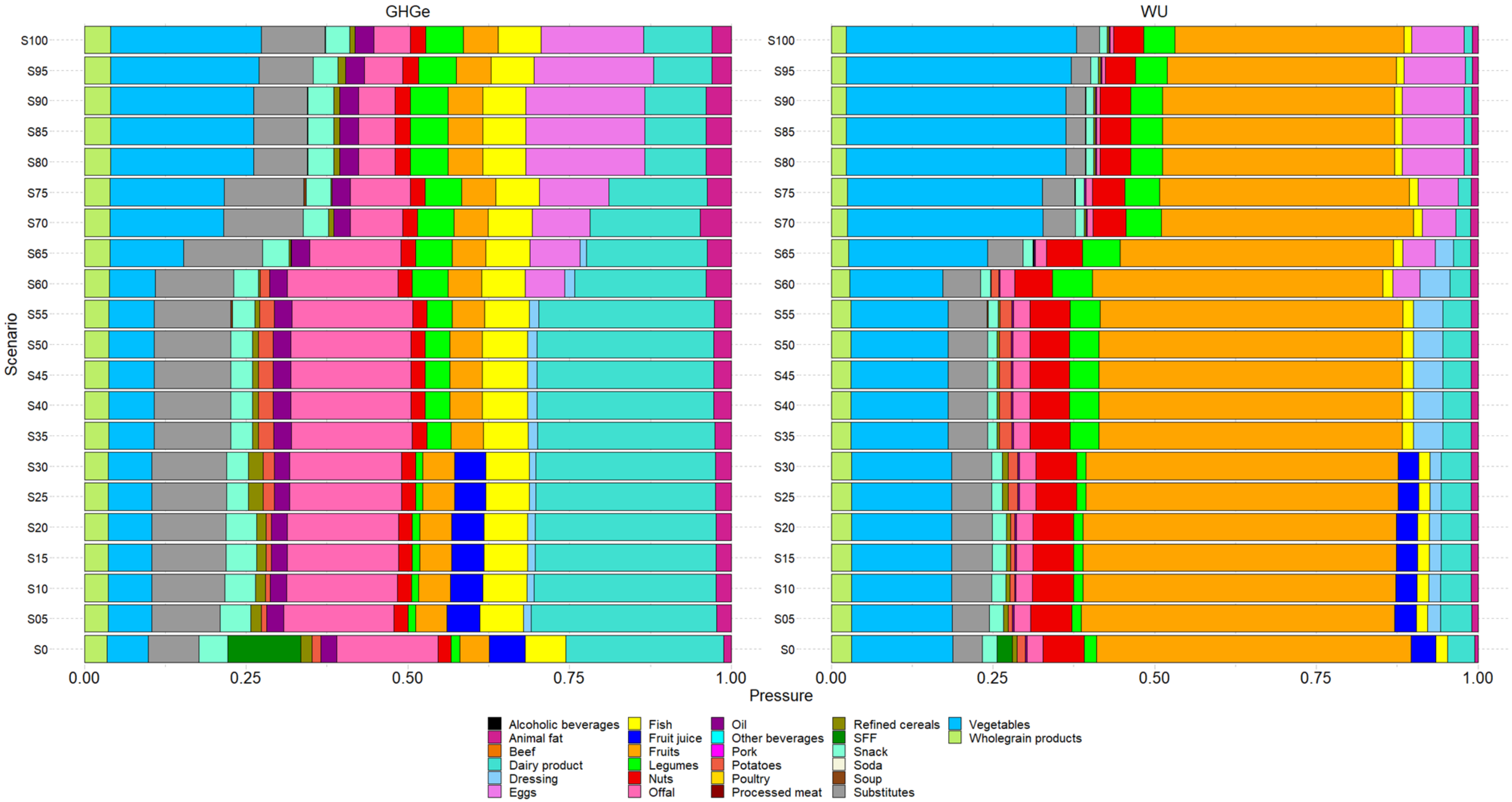

[1]Abbreviations: GHGe, greenhouse gas emissions (in kg eq $CO_2$); WU, water use (in eq world $m^3$/d). For clarity purpose, the 45 food categories were collapsed into 26 food groups

[2] Sλ denote the weight of GHGe in the objective function of the compromise modelling according the formula $Min\ OF = \lambda \times \frac{GHGe_i - GHGe_{best}}{GHGe_{worst} - GHEe_{best}} + (100\% - \lambda) \times \frac{WU_i - WU_{best}}{WU_{worst} - WU_{best}}$, λ=0% prioritization to water use reduction, λ=100% prioritization to GHGe reduction

**Supplemental Figure 2: Contribution of food groups to the Health Risk Score [1,2]**

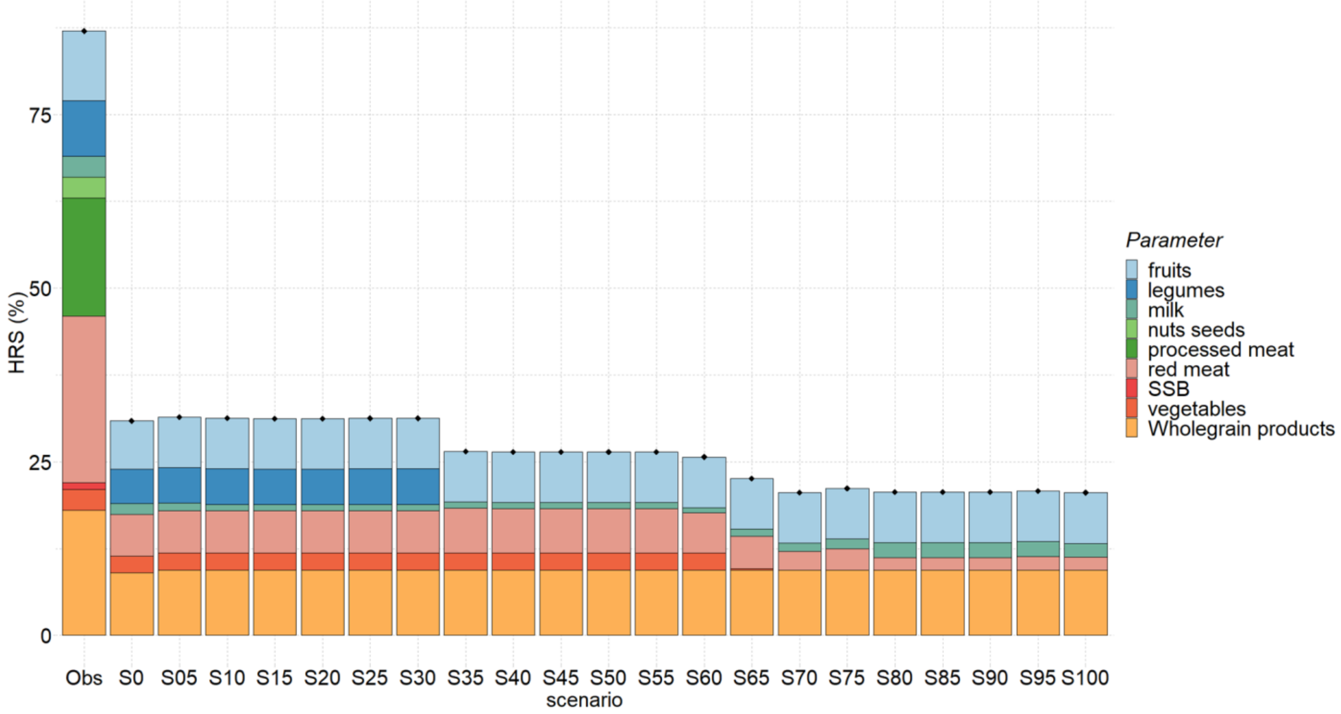

[1]Abbreviations: HRS, Health Risk Score; Obs, observed situation; SSB, sugar-sweetened beverages

[2] Sλ denote the weight of GHGe in the objective function of the compromise modelling according the formula $Min\ OF = \lambda \times \frac{GHGe_i - GHGe_{best}}{GHGe_{worst} - GHEe_{best}} + (100\% - \lambda) \times \frac{WU_i - WU_{best}}{WU_{worst} - WU_{best}}$, λ=0% prioritization to water use reduction, λ=100% prioritization to GHGe reduction

**Supplemental Figure 3: Food groups consumptions, GHGe and water use for the best-balanced modeled diet by sex[1]**

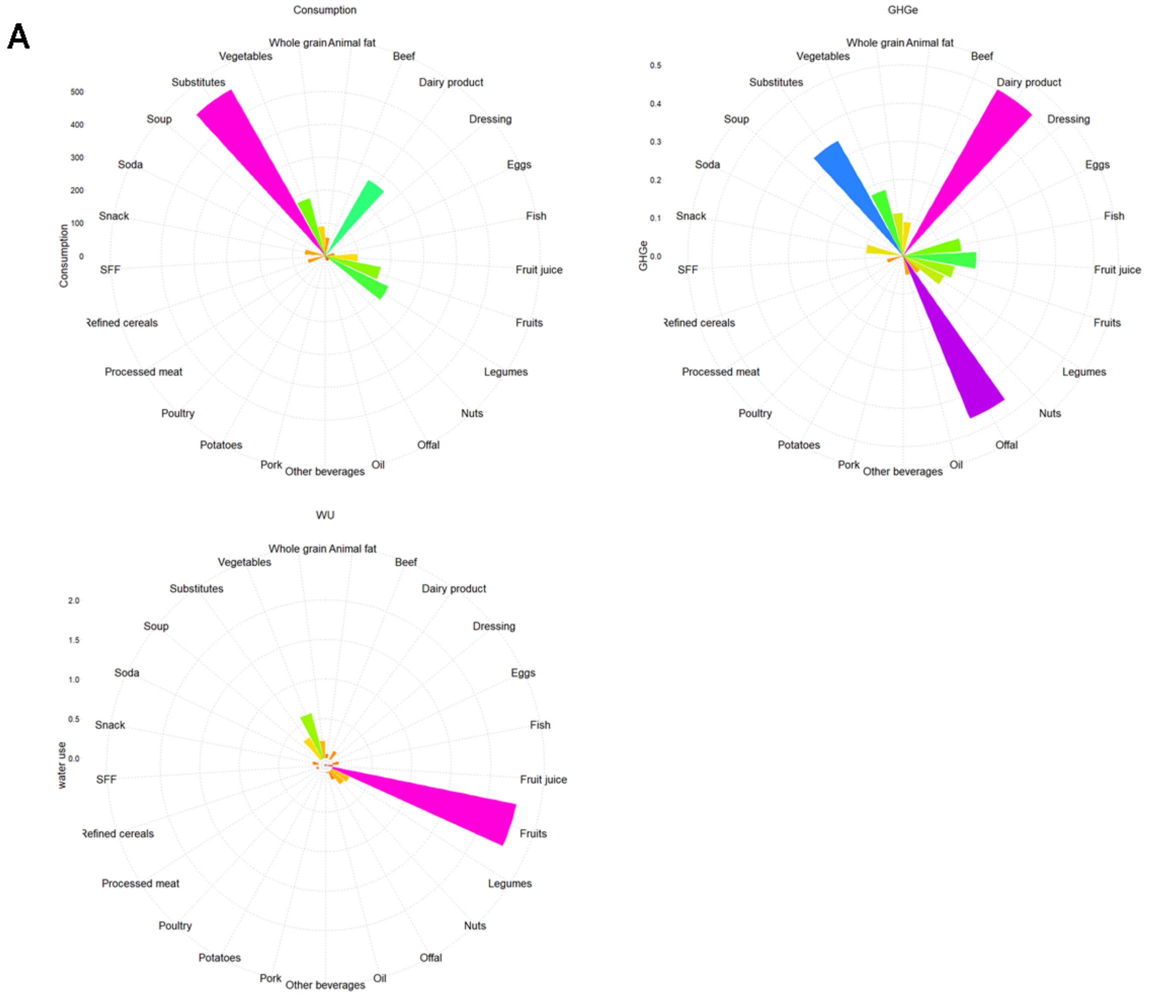

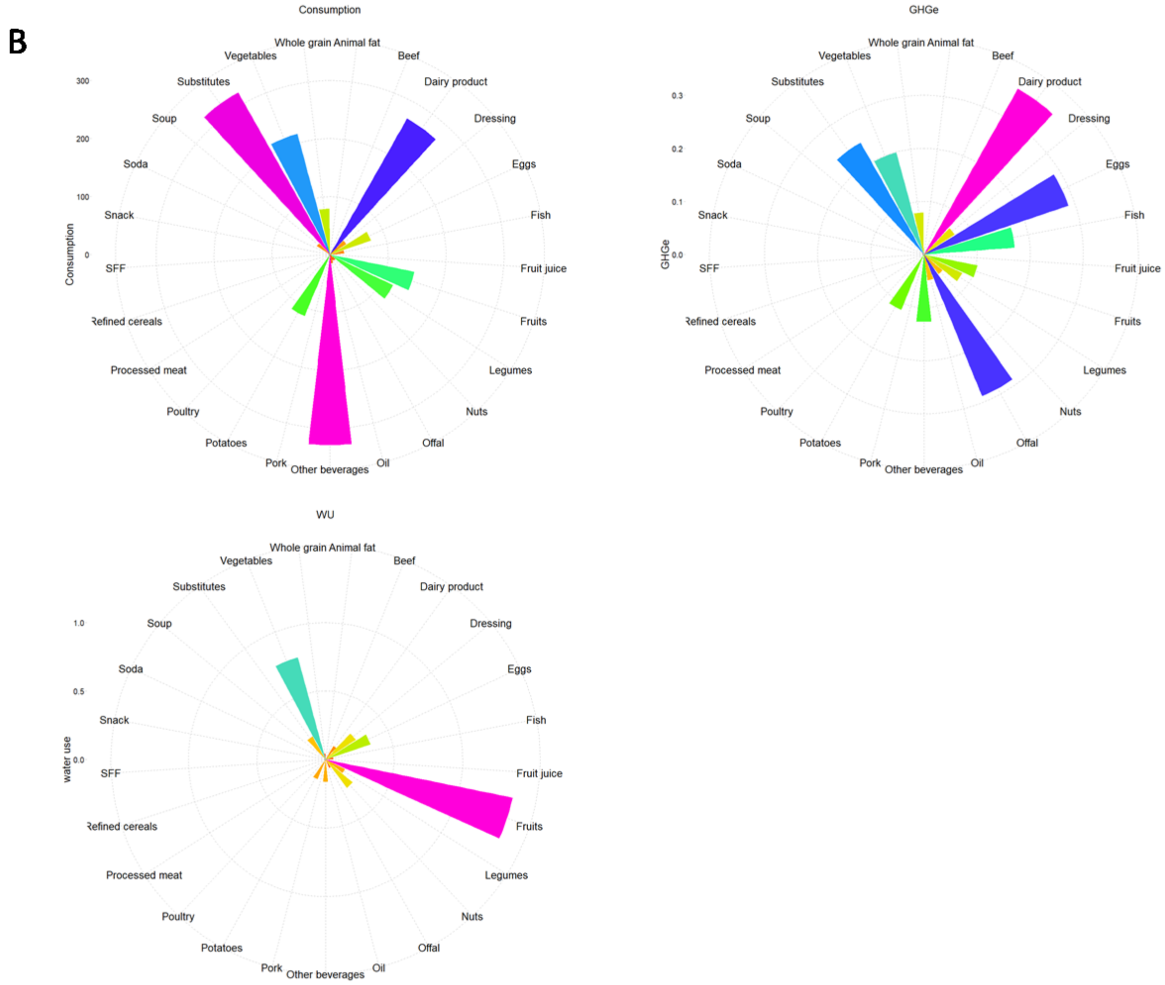

Abbreviations: GHGe, greenhouse gas emissions; WU: water use. For clarity purpose, the 45 food groups are pooled into 28 broader food categories.

[1] Food group consumptions in g/d, and contributions of food groups to GHGe in kg $CO_2$ eq /d and to WU in $m^3$ water eq deprivation /d for the best-balanced modeled diet identified in males (Panel A) and females (Panel B). The best-balanced modeled diet is the best compromise between efficiency and equity, identified by minimizing $(d_{GHGe} + d_{WU}) + \max(d_{GHGe}, d_{WU})$, where $d_{GHGe}$ and $d_{WU}$ are the normalized distances to the best GHGe and WU values, respectively.

**Supplemental Figure 4: Food groups consumptions, GHGe and water use for the best-balanced modeled diet identified when being stricter regarding cultural acceptability [1]**

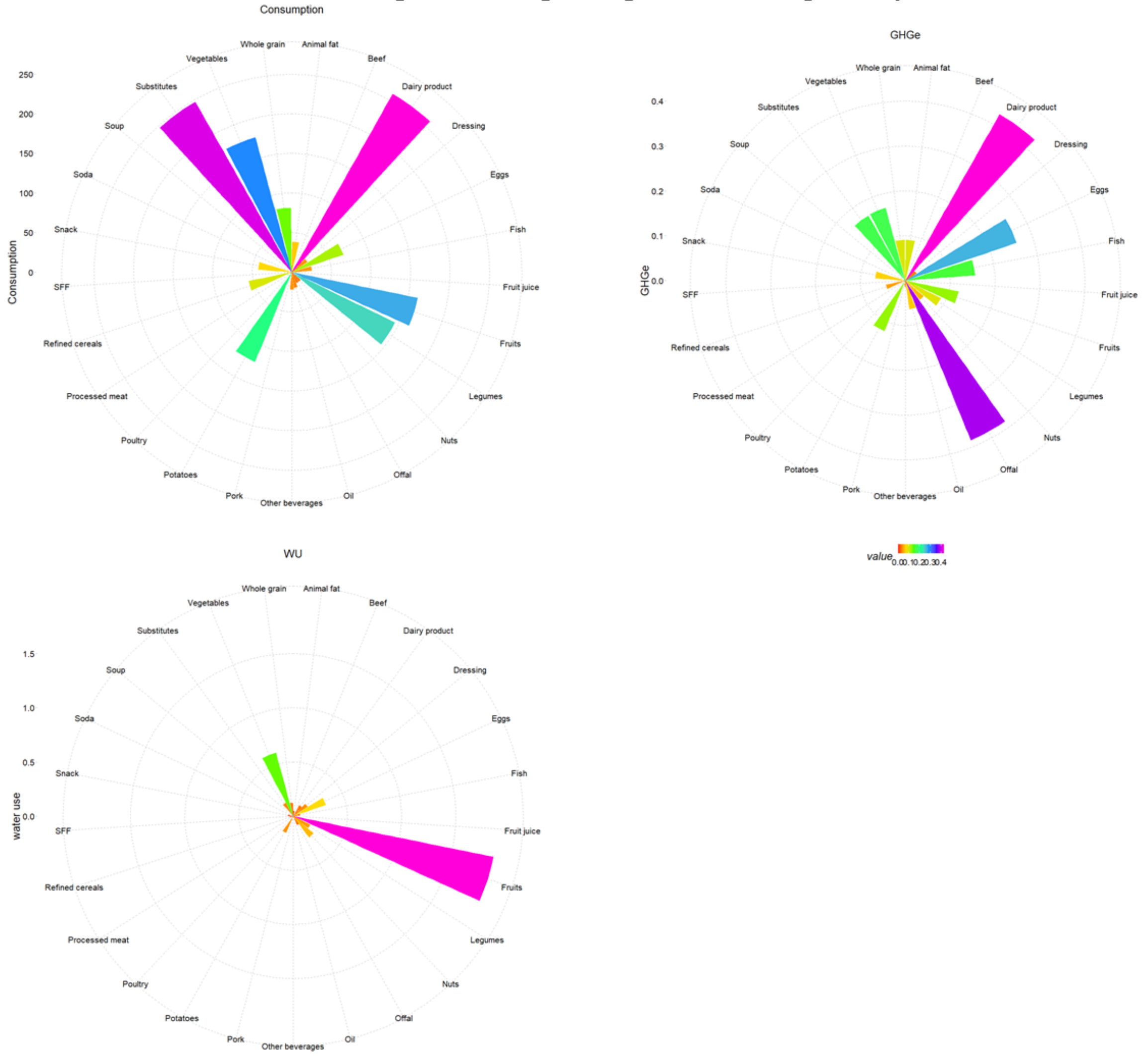

Abbreviations: GHGe, greenhouse gas emissions; WU: water use. For clarity purpose, the 45 food groups are pooled into 28 broader food categories.

Upper bounds for consumption initially set at the 99th percentile were modified at the 95th percentile (except for the vegetable oil rich in ALA needed to meet the nutritional constraints).

[1] Food group consumptions in g/d, and contributions of food groups to GHGe in kg $CO_2$ eq /d and to WU in $m^3$ water eq deprivation /d for the best-balanced modeled diet regarding efficiency and equity, identified by minimizing $(d_{GHGe} + d_{WU}) + \max(d_{GHGe}, d_{WU})$, where $d_{GHGe}$ and $d_{WU}$ are the normalized distances to the best GHGe and WU values, respectively.

**Supplemental Method 1: Identification of under-reporter for energy intake**

Energy under-reporting was identified using the method developed by Black's (1). It is based on the cut-off defined by Goldberg et al. (2), relying on the hypothesis that at stable weight, energy expenditure and intake are equivalent. Black's formulas are based on an estimate of the person's basal metabolic rate (BMR) calculated via Schofield's equations (3) considering sex, age, height and weight, and physical activity level (PAL), number of 24h records, as well as intra-individual variability of reported energy intake and BMR. In this study, intra-individual variation coefficients for BMR and PAL were fixed using the values proposed by Black et al., i.e. 8.5 % and 15%, respectively. To identify the under-reporters, the 1.55 value of PAL was used as recommended by WHO (4) and corresponding to the minimum energy requirement for a normally active sedentary individual. In this study 52 participants (corresponding to 12.55 % of the subjects aged under 65 years of age) were considered as under-reporters for energy intake and were excluded from the study.

**Supplemental Method 2: Computation of the Health Risk Score**

The IHME (Institute for Health Metrics and Evaluation) conducted the GBD (Global Burden Of Disease) study which aims to describe the morbidity and mortality of major diseases and health risk factors worldwide (9). It is the most comprehensive global observational epidemiological study to understand the health challenges facing people worldwide in the 21st century. The theoretical minimum-risk exposure levels (TMREL) defined in the GBD study refer to the levels of exposure that minimize the risk of death and chronic non-communicable diseases associated with a single dietary risk factor, i.e., the health risk associated to the over- or under-consumption of unhealthy or healthy food groups, respectively. The aim is to estimate recommended target intakes for each of these unhealthy and healthy food groups using an objective approach, rather than using expert opinions as in the conventional subjective approach. After analysis of all the available epidemiological data, the GBD study has defined TMREL values for 3 unhealthy food groups (red meat, processed meat and sweetened beverages) and 6 healthy food groups (wholegrain products, fruits, vegetables, legumes, nuts and seeds, and milk), and the GBD also provides the disability-adjusted life-years (DALYs) associated with the excessive or insufficient consumption of these unhealthy or healthy food groups in different countries.

Thereby, we have constructed a Health Risk Score (HRS) that evaluates, for a given diet, its global and normalized distance to all the targeted TMREL values (10), expressed in % (for similarity with the diet similarity score, DSI). HRS was thus designed to be minimal (HRS=0%) when the consumption of each unhealthy food group is lower or equal to its TMREL value and that of each healthy food group is greater or equal to its TMREL value. Inversely, HRS was designed to be maximal (HRS=100%) when the diet is at the maximal possible distance from each of the targeted TMREL values. Moreover, within HRS, the distance to each food group target is weighted by the relative importance of reaching this target compared to others in terms of DALYs, using the following formula:

$$\mathrm{HRS} = 100 * \left[ \sum_{i=1}^{3} \left\{ \frac{Cons\ (i)}{Max\ (i)} \times \frac{DALYs\ (i)}{DALYs\ (all)} \right\} + \sum_{j=1}^{6} \left\{ max \left( \frac{TMREL\ (j)\ -\ Cons\ (j)}{TMREL\ (j)};\ 0 \right) \times \frac{DALYs\ (j)}{DALYs\ (all)} \right\} \right]$$

*Where:*

- i: unhealthy food groups to be limited (red meat, processed meat et sweetened beverages)
- j: healthy food groups to be promoted (wholegrain products, fruits, vegetables, legumes, nuts and seeds, milk)
- Cons: consumption of the item i or j
- Max (i): upper limit of consumption of the food group i (g/d) during diet optimization
- TMREL (j): TMREL value for the food group j (g/d)
- DALYs (i), DALYs(j): DALYs associated with over- and under-consumptions of the food groups i and j respectively (years)
- DALYs (all): sum of all DALYs(i) and DALYs(j)

The TMREL and corresponding DALYs values used were the following:

| | | TMREL[1] (g/d) | | DALYs[2] (y) | |
|---|---|---|---|---|---|
| | | Males | Females | Males | Females |
| Healthy foods | Wholegrain products | 170 | 137 | 31 405 | 10 987 |
| | Fruits | 367 | 297 | 20 130 | 9 512 |
| | Legumes | 107 | 87 | 17 103 | 3 386 |
| | Vegetables | 339 | 274 | 9 342 | 3 090 |
| | Nuts and seeds | 16 | 13 | 6 531 | 1 355 |
| | Milk | 486 | 393 | 3 521 | 2 727 |
| Unhealthy foods | Red meat | 0 | 0 | 28 562 | 20 824 |
| | Processed meat | 0 | 0 | 14 346 | 6 288 |
| | Sweetened beverages | 0 | 0 | 4 105 | 1 791 |
| Total | | | | 135 045 | 59 961 |

[1]According to the most recent (2019) estimates from the GBD, the TMREL values are of 0 g/d for red meat, processed meat and sweetened beverages, and of 150, 325, 95, 300, 14.5 and 430 g/d respectively for wholegrain cereal products, fruits, legumes, vegetables, nuts and seeds, and milk. As these TMREL values are global estimates corresponding to a mean energy intake of 2,300 kcal (9), we used gender-specific values adapted to the particular energy intake of men and women in our French population (centered around 2600 kcal and 2100 kcal in men and women, respectively).

[2]We used the most recent (2019) French gender-specific DALYs values associated with excessive/insufficient consumptions of unhealthy/healthy foods, available from the Global Health Data Exchange website (http://ghdx.healthdata.org/gbd-results-tool). In the present study, males/females ratio was 50%:50%.

**Supplemental Method 3: Agribalyse® database**

The diet-related environmental pressures were estimated by indicators resulting in matching consumption with the French database Agribalyse® 3.1, a recent update of the previous version 3.0.1 developed by the French Agency for the Environment and Energy Management (ADEME) and allowing a matching with the CIQUAL French food composition table (11). In the 3.1. update, a few identified errors were corrected, and some methodological improvements and enhancements have been developed. These improvements were carried out by EVEA S.A.S. Cooperative in collaboration with Agribalyse ® partners: GIS REVALIM, ADEME, ITERG, CIRAD, ACTALIA, ANMF, as well as the firm GINGKO21. This version has been reviewed by GIS REVALIM.

Environmental indicator estimations are based on the method of Life Cycle Assessment (LCA) whose scope is "from field to plate". The perimeter of the indicators covers each process of the value chain: agricultural production, transport, processing, packaging, distribution and retailing, preparation at the consumer's and disposal of packaging and these processes have been split into two phases 1) production and 2) post-farm. Of note, losses and wastes (other than the non-edible parts) at home as well as transport from the retail to the household have not been considered and are thus limitations. Overall, the method is based on the international LCA standards: ISO 14040 (12) and ISO 14044(13), LEAP guidelines (14) and product environmental footprint (PEF) (15) and the finalized indicators are provided per kg of product and are detailed per process.

For the agricultural phase of plant products, all upstream processes (notably input production) except storage or drying are included except for ingredients used in the case of processed food. In the case of animal products, all operations including the phases of production, transport and storage of feed, fattening of animals, milking, construction and maintenance of buildings and machinery have been considered. The scope chosen is consistent with those defined in GESTIM (16) and ecoinvent® (17). In AGRIBALYSE ® v3.0.1, The LCI (life cycle inventory) data covered the period 2005-2009, except for perennial crops (2000-2010). The variety of production systems was considered by applying coefficients based on the share of systems in national production. The allocation rules are varied and are based on international recommendations as described by the ISO 14040/14044 standards (12,13). In particular, allocations have been developed in order to distribute organic nitrogen fertilizers and mineral fertilizers (P and K) between crop sequences. Biophysical allocations were used for animal production (milk versus meat). The biophysical models used for animal production and allocations by type of productions are presented in the full report (18) according to the reference AFNOR-BPX 30-323 (19)(AFNOR, 2011) and in compliance with the ISO 14044 standard (13) according to 3 rules in descending order: 1) avoid allocation, 2): biophysical allocation and 3) economic allocation.
A characterization method recommended by the European Commission (Environmental Footprint 3.0) translates the input and output flows of the inventory into impacts. For the background data (inputs in construction, raw materials, etc.) the ecoinvent® database is used to assess the indirect emissions (off-field emissions). The full methodology and methodological choices have been already described (18).

Agribalyse® update 3.1 includes methodological changes (carbon sequestration, $N_2O$, and $NH_3$ flows, OLCA-Pest model (20), correction of land use flows, correction of nitrogen fertilizer data), the integration of new LCIs, new data on food processing (21,22) and imported products (World Food LCA Database, version 3.5), and the deployment of updates of the databases used as input, notably Ecoinvent (version 3.8). All the modifications are described in a change report (23).

The transition from commodities to food as consumed introduced coefficients related to the edible part and economic allocations between co-products. The recipes were disaggregated into ingredients, for feasibility reasons, a threshold of 95% of the ingredients covered was used. Therefore, for some ingredients such as salt, additives and spices, impacts were not estimated. Similarly, for the origin of the ingredients, a threshold of 70% coverage was used followed by a standardization step. Concerning consumption phase, only preparation at consumer's home was considered and waste and lost were not estimated.
The whole methodology and methodological adoptions have been described elsewhere (24) and post-farm estimations are aligned with the PEF guidelines (15).

A total of 16 midpoint indicators were available: GHGe, WU, land use, ozone depletion, particulate matters, ionizing radiation (effect on human health), ecotoxicity, cancer toxicity (effect on human health), non-cancer toxicity (effect on human health), photochemical ozone formation (effect on human health), acidification, terrestrial eutrophication, freshwater eutrophication, marine eutrophication, resource use, minerals and metals and resource use, fossils, carcinogenic toxicity, non-carcinogenic toxicity, and one endpoint ecological footprint score (EF 3) aggregating these 16 midpoint indicators. The normalization and weighting factors considered in the calculation of the EF 3 score have been extensively described (15).
In the Agribalyse ® database, water footprint has been estimated using the guidelines of The Water Footprint Network (*The Water Footprint Assessment Manual Setting the Global standard, 2011*) and refers to blue water. An extensive description of the water footprint in AGRIBALYSE® is available at https://doc.agribalyse.fr/documentation/documentation-complete ("*Empreinte eau*").

In addition, the EF 3 (environmental footprint) single score including 2 further indicators related to human toxicity is provided.
Concerning the indicators for food as consumed, and according to the guidelines of the PEF method (15), a quality indicator (DQR) is provided and the Agribalyse ® 3.0 database has been reviewed and criticized by RIVM and GreenDelta as well as by French agricultural and agri-food technical institutes "Peter Koch Consulting".
The Agribalyse database was matched with the INCA 3 data as they have a common coding.
A perfect match was possible for 97% of the food items. For the remaining items we have made matches for similar foods. The imputation was made more often for food groups of the following food groups: fruit and vegetable juices, meat (except poultry), non-alcoholic soft drinks, condiments, herbs, spices and sauces, fish, vegetables, shellfish, cheese, cold cuts, sugar and sweeteners, offal, hot drinks, pasta, rice, wheat and other refined cereals, vegetable fats, nuts, seeds, and oleaginous fruits. A limitation is related to the indicators for fruit juices because such data in Agribalyse database are very poor. For a list of items (spicy potato Wedges, wheat sprouts, Vegetable schnitzel or soy steak, ready-to-eat meat or vegetable broth, and dehydrated smeat or vegetable broth) no matches could be made, and its items were deleted.
In particular, the synthetic environmental footprint score uses the European EF3.1 method in its unofficial interim version. The EF3.1 interim method for Agribalyse ® was built based on the EF 3.0 adapted method (version 9.4). The major changes to the EF 3.0 adapted method concern 4 impact categories (ecotoxicity freshwater; human toxicity, cancer; human toxicity, non-cancer, and climate change).
Regarding climate change, some characterization factors of the EF3.0 method indicator have been modified based on the 6th report (AR6) of the IPCC (25). The normalization factors for the 3 impact categories Ecotoxicity freshwater; Human toxicity, cancer; Human toxicity, non-cancer have been modified based on the JRC method note EF3.1 (26)
In addition, approximately 50 new products, including plant substitutes, have been added. Some seasonal and out-of-season fruits and vegetables were distinguished. Out-of-season products were used for industrial products. Water use for washing fruits and vegetables was not considered except for the new products modeled (or remodeled) for Agribalyse 3.1, when they included this step. This is a limitation of the database.

**Supplemental Method 4: Bioavailability of iron and zinc**

***Iron bioavailability***

For iron bioavailability, we considered heme and non-heme iron (5).

The rate of absorption for heme iron was calculated as (6):

$$\text{Log Absorption } (\%) = 1.9897 - 0.3092 \times \log(SF)$$

where SF is serum ferritin (μg/L). We considered a stringent situation by setting serum ferritin at 15 mg/L. The rate of absorption for non-heme iron was calculated as (7):

Ln (%)= 6.294 – 0.709 ln (SF)+ 0.119 ln (VitCI)+ 0.006 ln (MFP + 0.1)
-0.055 ln(T+0.1)-0.247 ln (PhyI)-0.137 ln(CaI) -0.083 ln (NHI)

Ln Absorption (%)= 6.294 – 0.709 ln (SF)+ 0.119 ln (VitCI)+ 0.006 ln
(MFP + 0.1) -0.055 ln (T+0.1) -0.247 ln (PhyI)-0.137 ln (CaI) -0.083 ln (NHI)

where SF is serum ferritin (μg/L) which was also set at 15 mg/L, VitCI is vitamin C intake (mg), MFP corresponds to consumption of meat, fish, and poultry (g), T is tea intake (as number of cups), PhyI is phytate intake (mg), CaI is calcium intake (mg), and NHI is non-heme iron intake (mg).

***Zinc bioavailability***

For zinc absorption, we used the equation developed and updated by Miller and al. (8)

$$BZ\ (mmol/d) = 0.5\left\{0.069\left(1+\frac{PhyI\,(1-0.017CI)}{0.44}\right)+0.084+ZI\,(1+0.012PI) - \sqrt{\left(\left(0.069(1+\frac{PhyI(1-0.017\,CI)}{0.44})+0.084+ZI\,(1+0.012PI\right)^{2}-4\times 0.084\,ZI\,(1+0.012PI)\right)}\right\}$$

Where BZ is bioavailable zinc and PhyI, CI, ZI, and PI are phytates, calcium, zinc and proteins daily intakes, respectively. All variables are in units of mmol/d except protein which is in g/d.
